# Developmental Deconvolution for Classification of Cancer Origin

**DOI:** 10.1101/2021.11.15.21266314

**Authors:** Enrico Moiso, Alexander Farahani, Hetal Marble, Austin Hendricks, Samuel Mildrum, Stuart Levine, Jochen Lennerz, Salil Garg

## Abstract

Cancer is a disease manifesting in abrogation of developmental programs, and malignancies are named based on their cell or tissue of origin. However, a systematic atlas of tumor origins is lacking. Here we map the single cell organogenesis of 56 developmental trajectories to the transcriptomes of over 10,000 tumors across 33 cancer types. We use this map to deconvolute individual tumors into their constituent developmental trajectories. Based on these deconvoluted developmental programs, we construct a Developmental Multilayer Perceptron (D-MLP) classifier that outputs cancer origin. The D-MLP classifier (ROC-AUC: 0.974 for top prediction) outperforms classification based on expression of either oncogenes or highly variable genes. We analyze tumors from patients with cancer of unknown primary (CUP), selecting the most difficult cases where extensive multimodal workup yielded no definitive tumor type. D-MLP revealed insights into developmental origins and diagnosis for most patient tumors. Our results provide a map of tumor developmental origins, provide a tool for diagnostic pathology, and suggest developmental classification may be a useful approach for otherwise unclassified patient tumors.

## Introduction

Diagnosis of malignancy relies on histopathological classification of tumor appearance, often alongside other features such as mutation profiling and clinical presentation. However, many tumors display a spectrum of heterogeneous appearances, which may in part reflect unknown differences in their development. Tumor heterogeneity can lead to diagnostic uncertainty, with either disagreement amongst pathologists, overdiagnosis, underdiagnosis, or inability to distinguish ‘gray zone’ cases between tumor types (1–4). Additionally, the precise cell type of origin for many cancers is unclear, and these malignancies are usually classified based on tissue of occurrence. This lack of diagnostic information contributes to treatment of many cancers using non-targeted therapies that have harsh toxicities and poor outcomes. Understanding cell types of origin or developmental pathways dysregulated in malignancies is a major goal in cancer biology, and would enable more precise diagnosis and therapeutic interventions targeted at these pathways. By definition, CUPs are of unknown origin, and represent a particularly deadly cancer with an average 5-year survival of 9-12 months (5,6). These observations argue for complete developmental maps of human tumors and necessitate diagnostic tools that utilize developmental information.

Machine learning classifiers have shown promise as new tools when applied to image processing in radiology and histopathology (7–10). However, image classifiers only detect visual features and are sometimes subject to artifacts (11–13). Classifiers that use molecular features, such as gene expression, have great potential to aid in diagnosis through capturing non-visual information, and recent approaches have demonstrated value in combining visual and molecular features for classification (14–16). However, gene expression classifiers have suffered from overfitting due to the high number of features (17–19), which results in poor predictive power on new datasets, or have selected small gene panels for measurement, which also reduces predictive power and leaves information on the table. How to reduce the number of features while extracting the most relevant information from gene expression data remains a key challenge in building integrated diagnostic models.

To address these challenges, we made use of two comprehensive atlases: The Cancer Genome Atlas (TCGA) and the Mouse Organogenesis Cell Atlas (MOCA) (20,21). TCGA contains high quality expression data for 33 bulk sequenced, tumor and normal tissue types accompanied by diagnostic annotations. MOCA, in turn, contains single cell expression profiling (scRNAseq) dissecting the mammalian organogenesis process during E9.5 to E13.5 post-fertilization in mouse (corresponding to E22 to E44 in humans), when most main adult mammalian tissue lineages are developed. MOCA cells are classified into 10 main and 56 sub-developmental trajectories on the basis of gene expression similarity and through known marker genes (21).

In this analysis we systematically compare both atlases to construct a developmental map of human tumors. We use this map to define a deep learning artificial intelligence tool that extracts the developmental trajectory of tumors based on gene expression data and predicts tumor types with up to 99% accuracy. We apply this tool to CUPs and make predictions for patient samples, narrowing the differential diagnosis and with implications for treatment.

## Results

### Systematic mapping of The Cancer Genome Atlas tumors by developmental trajectories

An overview of the systematic comparison made between TCGA and MOCA datasets and its use in constructing a diagnostic tool is given (Fig. 1). In brief, we mapped tumors to trajectories belonging to major cell lineages and developmental programs (Fig. 1A-B). This allowed us to deconvolute bulk tumor gene expression signatures into developmental components (Fig. 1C), which we then fed into a Multilayer Perceptron classifier that outputs tumor type (Fig. 1D).

**Figure 1:**
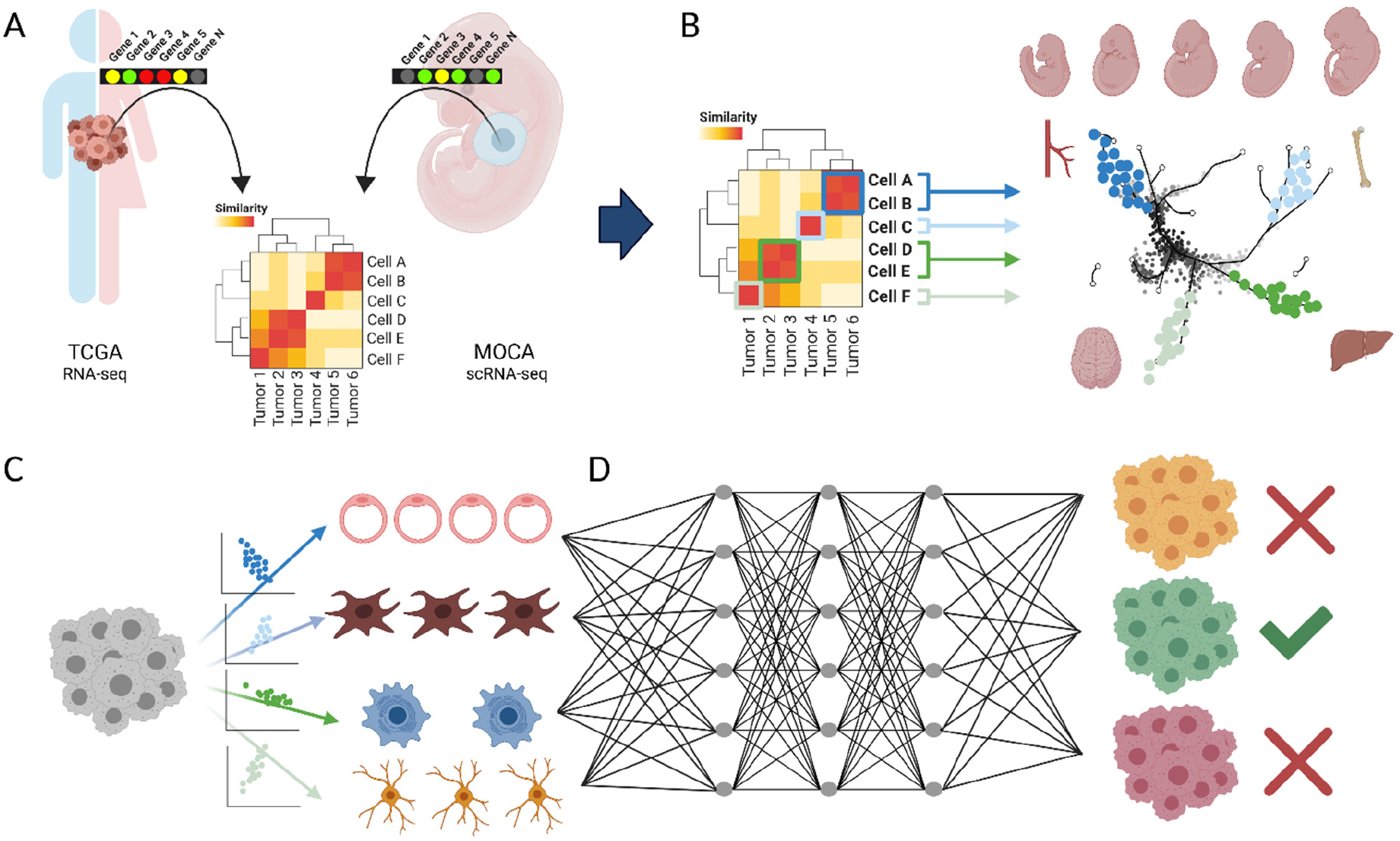
Diagnosis of Malignancy by Developmental Deconvolution and Machine Learning. **A.** A comparison between bulk RNA-sequencing data from The Cancer Genome Atlas (TCGA) and single cell RNA-sequencing data (scRNAseq) from the Mouse Organogenesis Cell Atlas (MOCA) was performed, generating a systematic developmental map of tumors. **B.** Each mapped relationship consists of tumor types and developmental sub-trajectories, represented at different stages of embryogenesis. **C**. Using this map, bulk gene expression signatures can be deconvoluted into component developmental trajectories that comprise each tumor sample. **D.** Scores for each developmental trajectory at each embryogenic timepoint can be fed into a Multilayer Perceptron classifier that examines these inputs and outputs tumor type prediction. This classifier is then applied to Cancer of Unknown Primary (CUP, not shown).

In order to achieve this goal, first we systematically compared gene expression profiles of each TCGA sample with each MOCA single cell. Altogether, the analysis constituted systematic expression comparison between 15,929 genes expressed across 10,388 TCGA samples derived from 9,681 patients (cohort details, Table S1) and 1,331,984 single cells derived from the MOCA dataset. In total, 21,217,199,458 data points were used to compute 13,836,649,792 correlation coefficients. Descriptive statistics on correlations are given (Table S2). To visualize the data, we utilized TCGA tumor type and MOCA cell sub-trajectories. Specifically, we plotted a matrix whereby every entry contained MOCA cells belonging to a specific developmental subtrajectory (rows) colored by each cell’s similarity with different TCGA tumor/tissue types (columns, i.e. primary, metastatic, or normal tissue) (Fig. 2A). Within each grid unit, MOCA single cells are plotted by their Uniform Manifold Approximation (UMAP) coordinates given by the MOCA dataset and colored by a single composite gene expression similarity signature for TCGA samples of each type. We examined the similarity between sample types and subtrajectories, noting many expected relationships. For example, inhibitory neuronal trajectories showed similarity with low grade gliomas (LGG) but not hepatocellular tumors (LIHC), and vice versa for hepatocyte trajectories (Fig. 2B).

**Figure 2:**
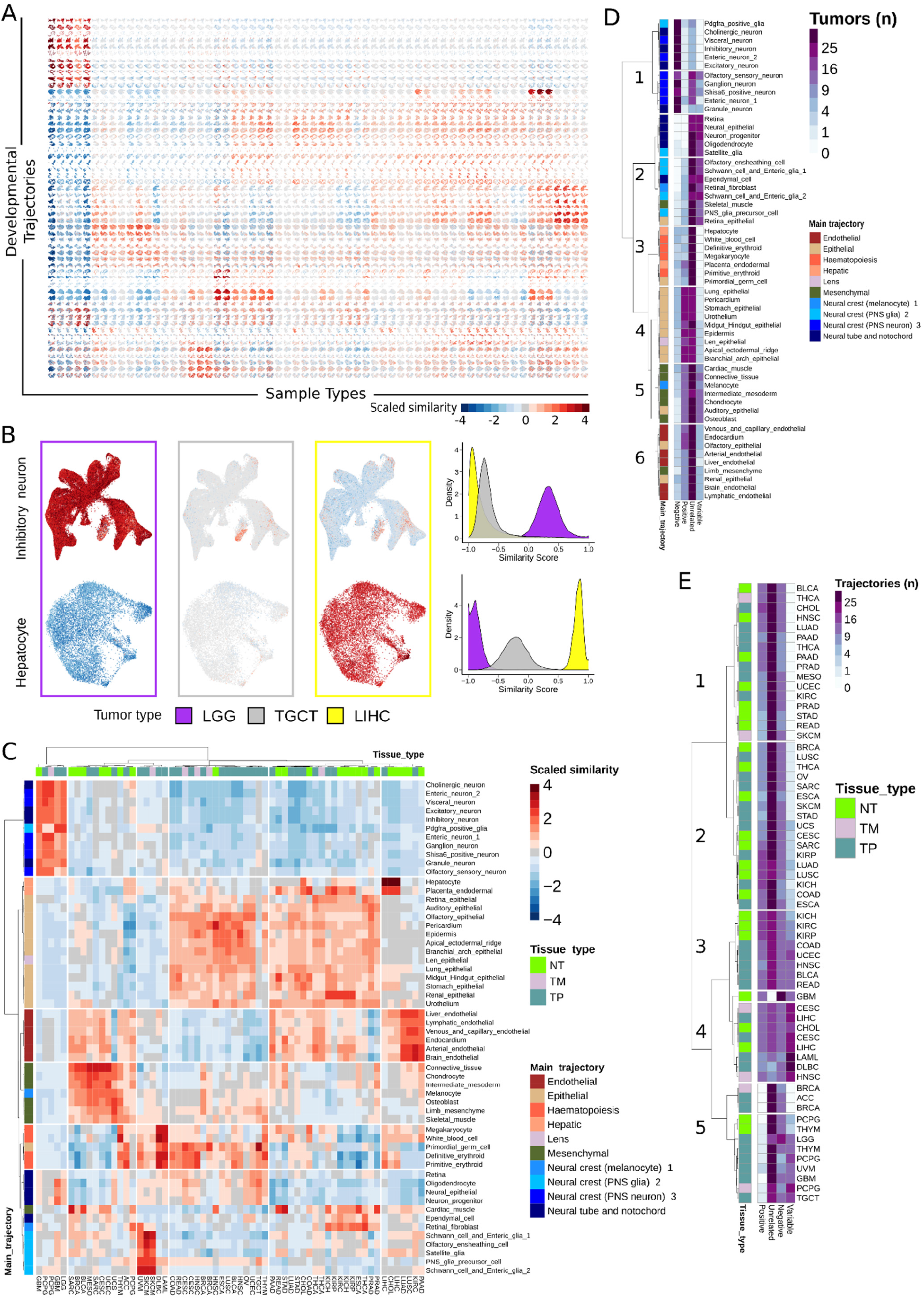
Systematic Mapping of TCGA to Developmental Trajectories. **A.** Each column contains 1,331,984 high confidence scRNAseq cells from the developmental atlas colored by similarity to each sample type in TCGA. Similarity was measured using Spearman’s rank-ordered correlation coefficient (ρ) for expression of each gene between each cell and each TCGA sample, and scaled across the dataset by Z-score (‘Scaled Similarity’, blue to red). **B. (Left)** From part A, higher magnification view of two developmental trajectories (inhibitory neuron, hepatocyte) across three tumors (LGG=low grade glioma, TGCT = testicular germ cell tumor, LIHC = liver hepatocellular carcinoma). Developmental trajectory cells were plotted using UMAP coordinates given by the MOCA dataset (relating each cell’s gene expression to the other cells), and colored by the aggregated gene expression profile of each TCGA sample type. **(Right)** Distribution of similarity scores across MOCA single cells for each tumor type. Inhibitory neuron trajectory cells showed higher similarity scores with LGG than do hepatocyte trajectory cells, and vice versa for LIHC. TGCT was not significantly related to either trajectory. **C.** Heatmap showing the scaled similarity between every developmental sub-trajectory and each TCGA sample type. TCGA samples from normal tissue (NT, green), metastatic tumors (TM, gray), and primary tumors (TP, aqua) are indicated at top. Main developmental trajectory types (given by the MOCA dataset) for each sub-trajectory are indicated at left, and developmental sub-trajectory names are listed at right. Hierarchical clustering of rows (trajectories) and columns (sample types) is shown. See Table S1 for TCGA sample codes. **D.** Each similarity relationship in part C was categorized as positive, negative, variable, or uninformative, and the number of TCGA tumor types falling into each category was summed and plotted for each developmental sub-trajectory. Hierarchical relationships are shown and selected clusters numbered for reference. **E**. As in D, but for TCGA sample types summating the number of developmental sub-trajectories falling into each similarity category.

Next, we collapsed correlation coefficients across all cells of the same developmental sub-trajectory and visualized them as a single similarity score (Fig. 2C). In other words, this heatmap scores similarity between TCGA samples aggregated by tissue type and MOCA cells aggregated by developmental sub-trajectory. Hierarchical clustering analysis of these data identified 6 TCGA sample and 6 developmental sub-trajectory clusters, for a total of 36 (Fig. 2C). Clusters highlighted broad relationships between tissue types and developmental trajectories: brain derived samples (LGG, GBM) with neuronal subtrajectories; kidney tumors (KICH, KIRP) with renal epithelial trajectories; hepatocellular tumors (LIHC) with hepatocyte trajectories; and testis tumors (TCGT) with germ cell trajectories, amongst several other expected correlations. Furthermore, we observed expected developmental lineage relationships: melanoma samples (SKCM) with neural crest trajectories, carcinoma tumors with epithelial lineages, and mesenchymal tumor types with mesoderm derived developmental lineages all showed strong similarity (Fig. 2C).

Comparisons between tumor and normal tissue types also revealed relevant biology (Fig. S1). Normal and malignant tissue types from similar anatomic locations were more likely to be clustered with each other, consistent with previous observations that TCGA tumor expression data largely reflects cell type of origin (22). In general, normal tissues tended to have higher scores for vascular endothelial trajectories, highlighting the abnormal vascularization associated with malignancy (23,24). Tumors characterized by strong inflammatory responses, such as bladder cancer (BLCA) and lung adenocarcinoma (LUAD), showed strong similarity with hematopoietic trajectories when compared to normal samples from the same tissues. Additionally, tumor-normal comparisons showed a relative loss of differentiation programs upon malignant transformation, such as a reduction in hepatocyte lineage similarity in cholangiocarcinoma (CHOL) compared to normal gallbladder (Fig. S1). Identification of expected developmental relationships and known tumor-normal relationships supported the notion that the observed correlations (Fig. 2A-C) were due to underlying biological relationships and served as partial validation of the approach. In order to further validate the observed correlations and approach, we developed an optimized protocol for transcriptome sequencing from formalin fixed paraffin embedded tissue (FFPE), sequenced the transcriptome for 40 tumors of various known types from FFPE, and compared results to TCGA trajectory correlations. Results showed excellent agreement (Fig. S2A, average Spearman ρ=0.69 between TCGA and FFPE cohorts), validating the overall approach.

Interestingly, comparisons of tumors to developmental trajectories also revealed unexpected or emerging relationships. Recently, some epithelial derived tumors such as lung adenocarcinoma have been noted to change their phenotypic characteristics in favor of parallel developmental pathways (25,26). The lung bud develops from the anterior foregut in embryonic week 3. We observed that lung derived tumors (LUAD and LUSC) showed strong similarity with gut derived trajectories, such as stomach and midgut/hindgut, and contrasted with normal lung tissue which did not show these similarities (Fig. 2C, Fig. S1). Therefore, this may represent re-expression of an earlier embryonic developmental program in these tumors. Additionally, glioblastomas represent a particularly heterogeneous tumor, with the exact cell type(s) of origin and their contributions to pathogenesis relatively unclear (27–30). Our analysis noted strong correlation of glioblastomas with main developmental trajectories neural tube/notochord and with a particular neural crest subtype, whereas other main trajectory neural crest lineages did not show such strong similarity (Fig. 2C). Thus, systematic comparison revealed many specific relationships between tumors and developmental trajectories with both expected and novel insights.

Next, we assessed which trajectories yielded the most information overall, by considering how often each trajectory was positively or negatively correlated with a tumor type. We also considered variable relationships, whereby a trajectory could contain cells with both high and low similarity to a tumor such that the net effect averaged out (Fig. S2B). We observed highly informative trajectories with strong positive and negative correlations (e.g. neuronal trajectories, Fig. 2D, cluster 1), trajectories with mixed relationships to tumors (e.g. endothelial trajectories, Fig. 2D, cluster 6), and many other relationships. A similar analysis was performed for each TCGA tumor type by examining the number of trajectories positively, negatively, and variably correlated with each (Fig. 2E). As expected, many TCGA tumor had trajectories with which they were positively and negatively correlated (Fig. 2E, clusters 1-2, 4-5). Interestingly, one group of tumors was characterized by anatomic location in the retroperitoneum (KICH, KIRC, KIRP, COAD, UCEC, BLCA, READ) and showed all types of correlative relationships (Fig. 2E, cluster 3). Together these analyses gave a strong map of TCGA sample types to developmental trajectories.

### Developmental time maps differences between tumors and normal tissue and is related to histological grade

Many cancers have been posited as diseases of dedifferentiation (31,32). The MOCA study dissected the organogenesis process in mice between E9.5 and E13.5 by examining multiple embryos at each timepoint. We made use of this by asking how relationships between samples and trajectories changed over time when comparing tumor and normal tissue, to assess if the former was dedifferentiated compared to the latter. We plotted the systematic correlations for normal tissue next to its corresponding tumor across a developmental trajectory while also noting the embryonic day for each cell (Fig. 3A). Glioblastoma shows a high degree of similarity with neuronal cells from all timepoints, whereas normal brain is more similar to neural cells from later timepoints (Fig. 3B, p < 2*10^−16^ for all timepoints except E12.5). Next, we extended this analysis to a pan-cancer cohort comprising all TCGA tumor types for which normal and primary tumors were available (Table S3). We identified the MOCA cells with highest positive correlation to each sample and binned the known embryonic time period of these cells. This allowed us to compare tumor samples to normal samples using categoric enrichment chi-square testing (χ^2^). We found a pan-cancer enrichment for earlier embryonic periods in tumors compared to normal tissue (Fig. 3C, p < 2.22*10^−16^, χ2 test). Further, we computed a normalized ‘embryonic period score’ for each TCGA sample (Methods). Tumors were shifted towards a lower embryonic period score (Fig. 3D, p < 1.4*10^−119^ K-S test), consistent with the idea that malignancy is driven in part by dedifferentiation.

**Figure 3:**
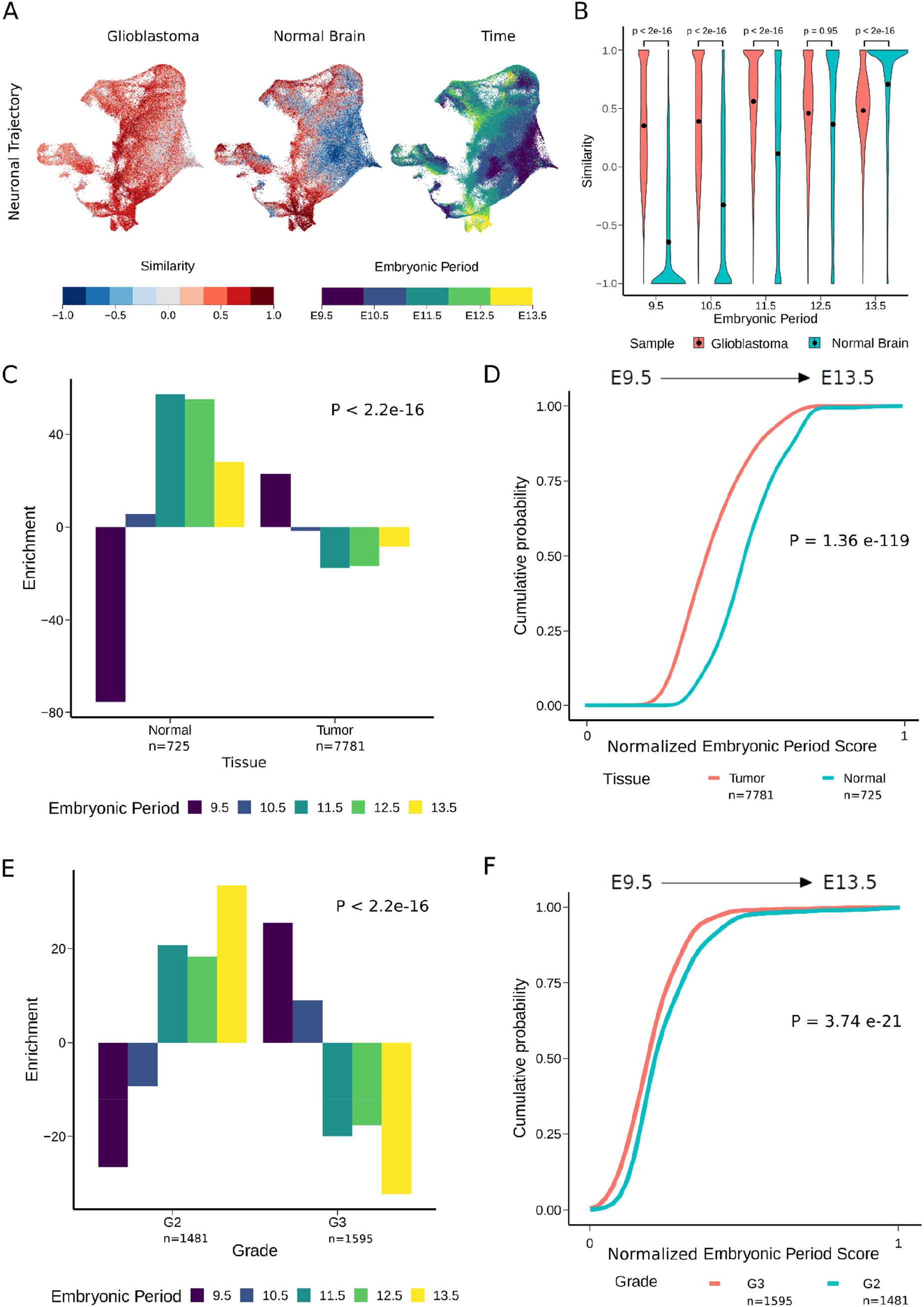
Validation Through Comparing Relationships Between Sample Types and Developmental Time. **A**. Representation of the similarity of neuronal trajectory cells compared to both glioblastoma and normal brain samples is shown. At right, the developmental embryonic timepoint from which the cell was isolated is shown. **B.** Violin plots showing the distribution of similarity scores for each sample type (glioblastoma, normal) at each embryonic timepoint (E9.5 – E13.5), dots indicate the median cell. P-values for Mann-Whitney U-tests at each timepoint are shown. **C.** Pan-cancer comparison of tumor versus normal sample types for enrichment of their most similar developmental cells (Methods) at each embryonic timepoint. P-value for chi-square categoric enrichment testing is shown. **D**. Cumulative distribution (CDF) plot of normalized embryonic period score (see Methods) for all tumor and normal samples in TCGA is shown. P-value for Kolmogorov-Smirnov testing is shown. **E.** Pan-cancer comparison of grade 2 versus grade 3 tumors for enrichment of their most similar developmental cells at each embryonic timepoint. P-value for chi-square categoric enrichment testing is shown. **F**. CDF plot of normalized embryonic period score for grade 2 versus grade 3 tumors. P-value for Kolmogorov-Smirnov testing is shown.

Tumor grade is the assessment of histological characteristics of cancer and is related to malignant potential, whereby lower-grade tumors are more similar to normal tissue and higher-grade tumors exhibit significant architectural disarray. Analogously, higher grade tumors are posited to be more dedifferentiated than lower grade tumors. To assess this hypothesis, we analyzed whether tumor grade corresponded to developmental time, focusing on Grade 2 and Grade 3 tumors due to the availability of samples annotated at these grades in TCGA. A list of tumors analyzed is given (Table S3). Grade 2 tumors showed a relative enrichment for later embryonic periods compared to Grade 3 tumors (Fig. 3E, p < 2.22*10^−16^, χ2 test). Also, Grade 2 tumors were shifted towards higher embryonic period scores, consistent with greater similarity with later developmental stages (Fig. 3F, p < 3.74*10^−21^, K-S test). Together, the results suggested developmental mapping captured pertinent histopathological features of malignancy. Additionally, pan-cancer confirmation that tumors represented dedifferentiation further validated the biological underpinnings of the calculated correlations.

### Deconvolution of tumors into component developmental trajectories

The creation of a systematic map between TCGA samples and developmental trajectories inspired us to attempt a systematic developmental deconvolution (DD) of human tumor gene expression. In deconvolution, a recorded signal (gene expression) made of component parts (developmental programs) is deconstructed into individual signals from each component (trajectories at embryonic timepoints). We used developmental components (DC), a single quantitative measure of each developmental sub-trajectory at each timepoint, to represent the deconvolution signature for every TCGA sample (Table S4). DCs were scaled across all tumor samples and charted on radar plots, which represent information about developmental period, sub-trajectory, and DC score for each sample. A schematic for this plot is shown (Fig. 4A and Fig. S3). A representative radar plot for developmental deconvolution of a single hepatocellular carcinoma (LIHC) is shown (Fig. 4B). In this sample, we appreciated a strong enrichment in signal for hepatic trajectories (peach, 6 o’clock), depletion of neuronal trajectories (blue, 1 o’clock to 4 o’clock), limited signal for hematopoietic and endothelial trajectories (orange, 8 o’clock and maroon, 11 o’clock, respectively) and little signal for mesenchymal trajectories (green, 5 o’clock), in a pattern that extended to additional hepatocellular carcinoma samples (Fig. S4). However, differences were also noted amongst LIHC samples, particularly in deconvolution scores for endothelial trajectories, perhaps reflecting differing degrees of tumor vascularization (Fig. S4).

**Figure 4:**
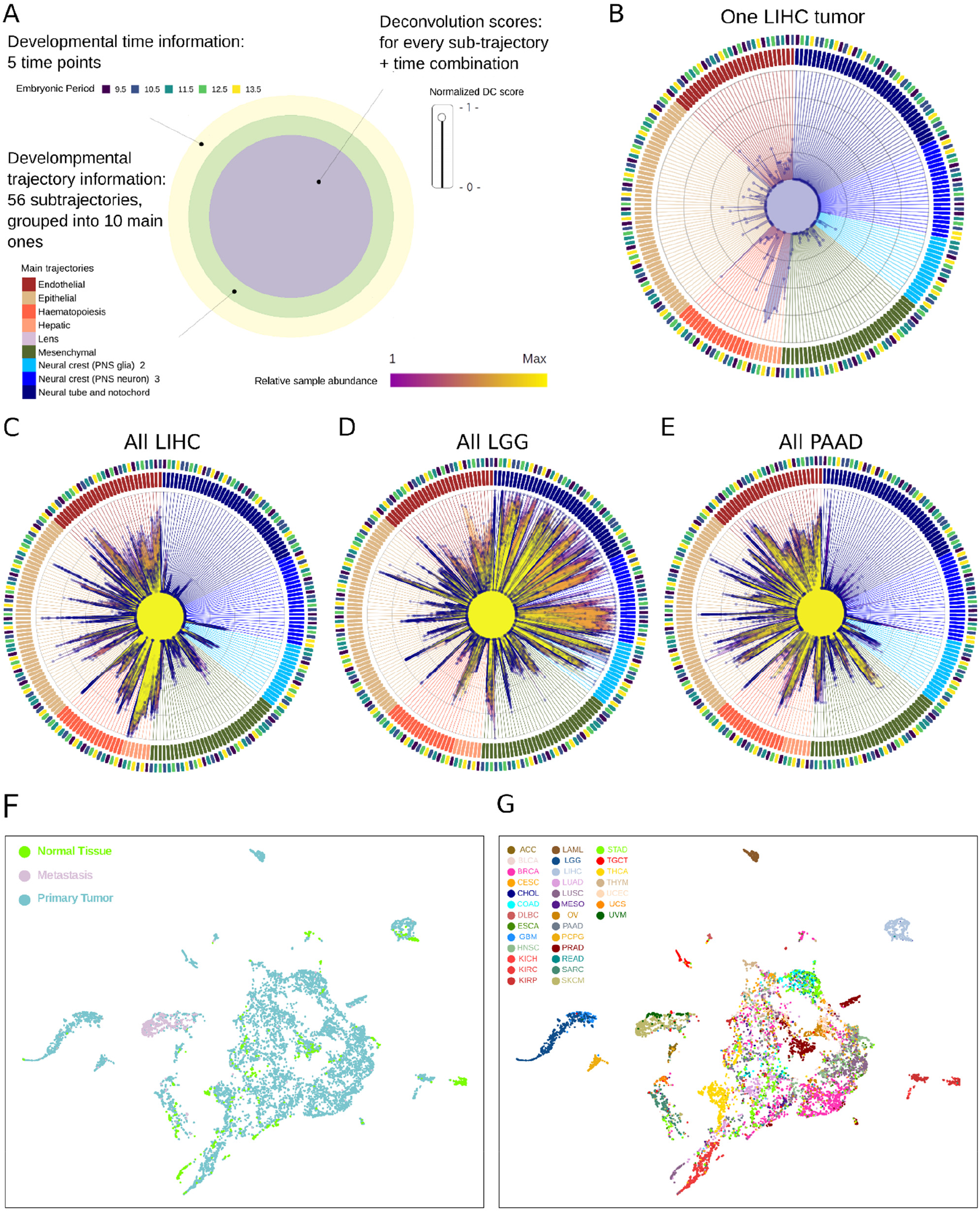
Developmental Deconvolution of Tumor Samples. **A.** Diagram of radar plots showing the results of deconvolution. Multiple layers of information are present. For each sample, a deconvolution score is generated for each developmental sub-trajectory at each timepoint (214 scores total), represented as the distance from center in the innermost circle. Timepoints and the main developmental trajectories are shown in the outer two circles. See also Fig. S3 for sub-trajectory order on the plot. **B**. Radar plot showing the developmental deconvolution of a single hepatocellular carcinoma sample (TCGA-DD-A1EB). **C**. Radar plot showing trajectory deconvolution signals for all hepatocellular carcinoma samples. The relative abundance of each trajectory across all tumor samples is shown by shading (yellow to magenta). See also Fig. S4. **D**. Radar plot showing trajectory deconvolution signals for all low-grade glioma samples. See also Fig. S5. **E**. Radar plot showing trajectory deconvolution signals for all pancreatic adenocarcinoma (PAAD) samples. **F**. UMAP dimensionality reduction was performed on the 214 developmental deconvolution scores for each TCGA sample. Sample type (normal, primary tumor, metastasis) is indicated. **G**. Identical to **F** but each sample is colored by tissue type. See also Fig. S8 for each tumor type colored separately.

We plotted the signal across all hepatocellular carcinomas in a single radar plot (Fig. 4C). All LIHC samples were characterized by elevated scores in hepatic trajectories and depletion in neuronal trajectories. In contrast, low grade gliomas (LGG, Fig. 4D) showed high signal for neuronal trajectories and low signal for hepatic trajectories, consistent with results from mapping (Fig. 2). Interestingly, the degree of sample-to-sample variation in endothelial DCs was lower for low grade gliomas than for hepatocellular tumors (Fig. S5–6), perhaps reflecting that these tumors are not well vascularized. However, LGG tumors did show variable deconvolution into neuronal trajectories, which may reflect differences in anatomic location in the brain from which each was isolated, or also could reflect patient heterogeneity in the precise developmental context in which each tumor arose. Other tumor types also showed tumor specific patterns, such as pancreatic adenocarcinomas (PAAD), which had high DC scores in epithelial trajectories (Fig. 4E), as expected. The spectrum of radar plots for each tumor type is shown (Fig. S7) for comparison.

Next, we analyzed the effectiveness of developmental deconvolution at separating different tumor types from one another. We plotted developmental component scores across all TCGA samples using UMAP dimensional reduction (214 DC scores inputted per sample, Methods), annotating by tissue type (normal, primary tumor, metastases, Fig. 4F) and by tumor type (TCGA code, Fig. 4G and Fig. S8). Normal tissue and primary tumor were spread throughout the plot, and clustered largely according to tissue of origin. Further, most tumor types were strongly clustered (e.g., LIHC, light gray and LGG, medium blue, Fig. 4G and Fig. S8), confirming that developmental deconvolution extracted salient gene expression data. However, overlaps were also noted, with some tumor types (BRCA, breast adenocarcinoma, magenta) spread throughout. Metastatic samples clustered together (Fig. 4G), partially due to the overrepresentation of cutaneous melanoma (SKCM) in this group. Together, this suggested the TCGA-MOCA mapping could be used to deconvolute tumor gene expression into developmental components, in a manner that resolved most tumor types.

### Construction of the Developmental Multilayer Perceptron (D-MLP) classifier for cancer type prediction

The ability to resolve different tumor types by developmental deconvolution raised the possibility of designing a supervised machine learning (SML) approach to classify malignancies. SML approaches have been previously applied to gene expression data but have been limited by difficulties with model overfitting and representation, due to the high dimensionality of the transcriptome (∼22,000 protein coding genes) (33,34). To avoid these issues, some studies have selected small gene subsets (35–37), but this restricts model input to selected datapoints and compromises accuracy and predictive power. We reasoned a classifier based on developmental deconvolution scores would extract most relevant data from gene expression (Fig. 5A), while also capturing non-linear relationships between genes in the form of embryological development programs dysregulated in tumors. After literature mining (38) and testing different approaches, we decided on a Multilayer Perceptron due to its ability to simultaneously perform feature extraction and classification on a dataset. This class of SML algorithms relies on artificial neurons, or threshold logic units, organized in three classes of layers (input, hidden and output) and takes advantage of back-propagation to increase its classification accuracy.

**Figure 5.**
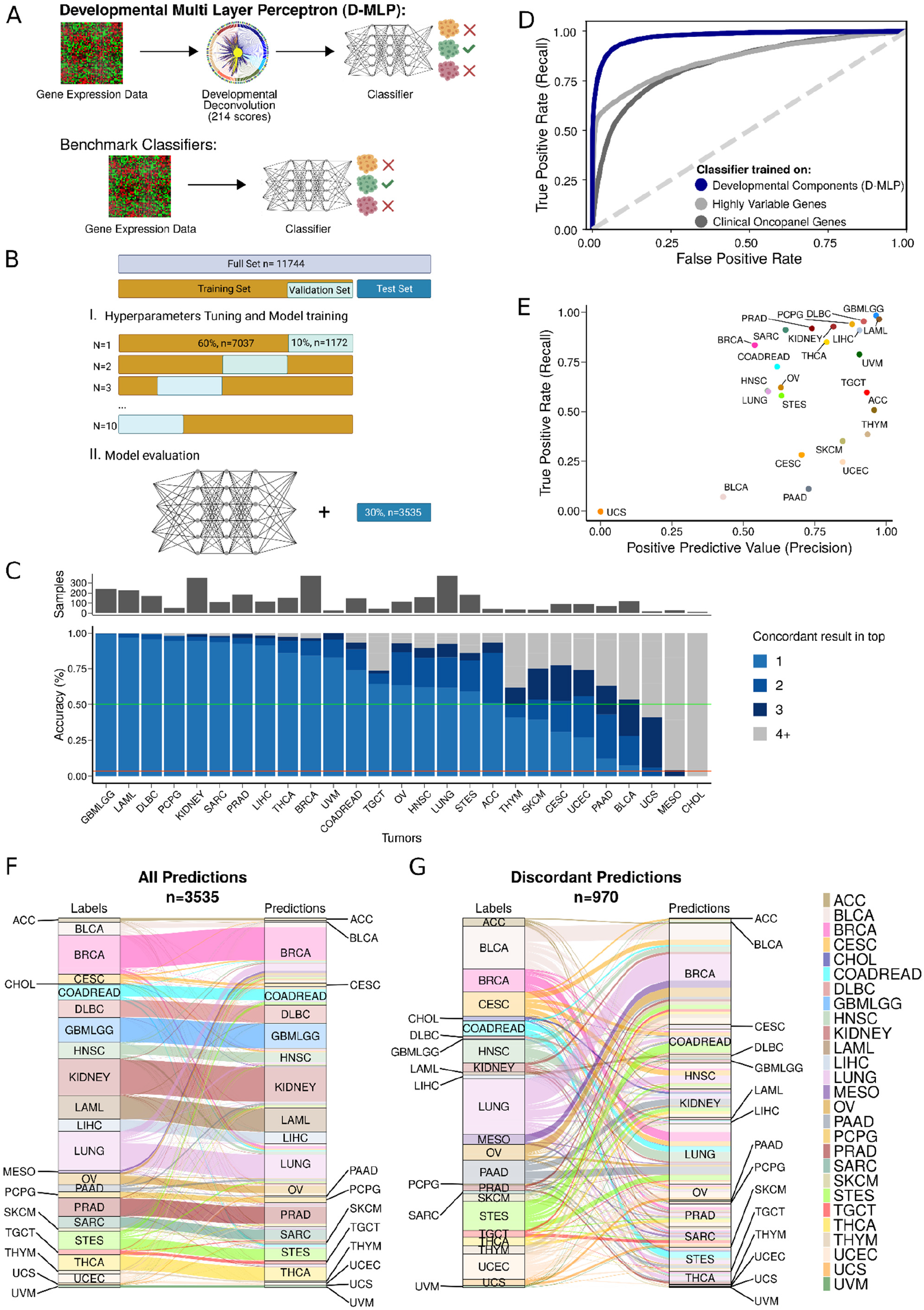
Construction and Testing of Developmental Multilayer Perceptron (D-MLP) Classifier for Tumor Type. **A.** Schematic for classifier construction and testing. The D-MLP classifier uses developmental deconvolution scores calculated as the similarity of each tumor’s gene expression to embryologic developmental trajectories (Fig. 4) as input. Comparison is made against benchmark classifiers that use gene expression data directly. **B.** Parameter optimization and model training. Of the full cohort (11,744 samples), **(i)** 70% of cases were sampled, 60% for training and 10% for validation, in hyper-parameter optimization using a 10-fold cross validation approach to construct the classifier (see Methods for further details). **(ii)** 30% of cases were held out and never seen by the model during training or optimization (Test set). The model was assessed on these cases to gauge performance. **C**. Classifier accuracy (concordance) measured against the test set and number of samples are shown for all TCGA tumor types. **D**. Micro-averaged receiver operator curve (ROC) plotting the true positive rate (also known as recall) as a function of false positive rate for classifier performance for the top prediction. D-MLP classifier performance (blue line) and random guess performance (dashed grey line) are shown. Area under the curve (ROC-AUC) for the top prediction was calculated as 0.974 +/− 0.003. Also shown are ROC performance curves for benchmark classifiers trained on either the most highly variable genes in expression across the training cohort (‘highly variable genes’, light grey, ROC-AUC: 0.859 (95% confidence interval 0.829 - 0.976) or expression of a panel of genes tested in routine clinical assays at our institution (‘clinical oncopanel genes’, dark grey, ROC-AUC: 0.836 (95% confidence interval 0.828 - 0.975). **E**. Precision (positive predictive value) versus recall (true positive rate) performance characteristics for each TCGA tumor type. Note CHOL and MESO are omitted as the positive predictive value is undefined for these tumors. **F**. Sankey plot showing classifier results for the top tumor type prediction for all samples (n=3,535). **G**. Sankey plot showing the results for discordant classifications (n=1,006) for top predictions.

First, we assembled a broad set of data. In addition to TCGA, we incorporated samples from other cancer cohorts (BEATAML1.0, CGCI-BLGSP, CTSP-DLBCL1, MMRF-COMPASS and TARGET) (5,39–41), and sequencing data we generated from FFPE tissue (Fig. S2A). FFPE samples are part of routine Pathology workflows; their inclusion allowed for a classifier more proximal to clinical implementation. To incorporate different studies together, we merged sub-classes for specific tumors together into main classes (Methods), leaving 27 diagnostic categories. This enlarged cohort (11,744 samples, Table S5) allowed us to increase sample size and build a model using data gathered by different methods. Next, we divided samples into two separate cohorts: a training cohort (70% of total, n=8,209) used to construct the model, and a separate cohort (30% of total, n=3,535) that was never seen by the model during training or optimization and was used later to test performance (Fig. 5B). After dividing the dataset, we used the training cohort for hyperparameter optimization, using a grid search and 10-fold cross-validation approach (see Methods). The final architecture was trained and validated ten times, each time drawing only from the 70% training cohort (60%, n=7,037 for training with 10%, n=1,172 for validation each iteration, Fig. 5B).

When trained on developmental deconvolution scores (214 per sample), the D-MLP classifier reached an overall “top1” prediction concordance of 73% and “top3” prediction concordance of 90% (Fig. 5C). For many tumor categories, (e.g. GBMLGG, LAML, DLBC, PCPG, KIDNEY, SARC) concordance rates of 92-100% were observed (Fig. 5C). Interestingly, BRCA tumors showed relatively high concordance despite being spread across developmental deconvolution (Fig. S8), further suggesting the D-MLP classifier was capturing relevant information. Overall, 24 out of 27 tumor types were classified with a higher degree of concordance than by chance. Lower concordance rates were obtained for some tumors (e.g., COADREAD, HNSC, LUNG, and STES), and very poor results for CHOL (cholangiocarcinoma), UCS (uterine carcinosarcoma), and MESO (Mesothelioma) (Fig. 5C and Fig. S9). Poor CHOL classification may be due to the lack of development of a distinct gallbladder in rodents (42), with no trajectory for this cell type in the MOCA data, poor UCS classification due to few training samples, and poor MESO classification due to the fact that mesothelioma arises from toxin exposure (asbestos) and may lack a developmental signature. Receiver operator curve (ROC) characteristics for classifier performance were strong, with a ROC area under the curve (ROC-AUC) of 0.9740 (micro-averaged method for top prediction, 95% confidence interval 0.971 – 0.977, Fig. 5D), and with high precision (positive predictive value) and recall (true positive rate) for most tumor types (Fig. 5E), together validating D-MLP effectiveness.

Next, we compared our developmental deconvolution approach to training directly on gene expression data. First, we selected the most variably expressed genes across our assembled case cohort. We used the 214 most variably expressed genes to match features counts with D-MLP, trained a new classifier using the same architecture and hyperparameter optimization approach, and evaluated accuracy of the new classifier on the test set. This classifier (‘highly variable genes’) performed with a ROC-AUC of 0.859 (95% confidence interval 0.829 - 0.976, Fig. 5C and S10), substantially under D-MLP performance. Second, we selected a panel of 214 genes tested in diagnostic clinical assays at our institution (e.g. EGFR, MYC, KRAS, amongst others, see Methods for details), trained a new classifier on expression of these genes using the same architecture and approach as D-MLP, and evaluated its performance on the test set. This classifier (‘clinical oncopanel genes’) performed with a ROC-AUC of 0.836 (95% confidence interval 0.828 - 0.975, Fig. 5C and S10), again under the performance of D-MLP. Additionally, these benchmark classifier approaches had lower overall accuracy (Fig. S10). We conclude developmental deconvolution yields higher accuracy classification than directly training on gene expression data for similar numbers of input features.

### Analysis of discordant D-MLP predictions

In principle, discordant predictions could arise from either classifier inaccuracy or from additional developmental information captured in tumor sequencing. Tumor sampling could include nearby tissue, tumors might share previously unappreciated developmental connections, or tumors classified as one histopathological entity might contain heterogeneity between samples in their developmental origins. To assess these possibilities, we examined discordant classification relationships amongst tumors (Fig. 5F-G and Fig. S9). The adrenal gland rests on top of the kidney within the retroperitoneum. We noted that adrenocortical carcinoma (ACC) was often discordantly classified as kidney (12%) or as an adrenal medulla tumor (9%, PCPG). Other examples pointed to tumor heterogeneity. For example, both lung and breast adenocarcinomas are epithelial tumors, both normal tissues are formed by the interaction of ectodermal and mesodermal elements, and both normal tissues continue remodeling into young adulthood (43,44). We found breast and lung tumors were commonly discordantly classified as each other (Fig. 5F), and shared striking heterogeneity in their signal for epidermis and branchial arch developmental trajectories across samples (Fig. S11). A summary of tumor types and trajectories amongst discordantly classified samples is shown (Fig. S12). In general, concordant predictions reflected known cancer biology while discordant ones reflected less well understood connections between tumor types or developmental heterogeneity within tumors.

### Classification of cancer of unknown primary

Cancer of unknown primary remains a major clinical problem, with malignancies that show aggressive features and poor patient outcomes. Of patients that present in clinic with CUP, a fraction are resolved upon routine pathological examination using hematoxylin and eosin staining (H&E), a process that can be aided by machine learning tools that rely on image inputs (9). An additional fraction of cases can be resolved using immunohistochemistry (IHC) and tumor mutation profiling (Fig. 6A). However, a portion of cases fail all currently available diagnostic techniques and remain true diagnostic dilemmas in need of new approaches (Fig. 6A, far right). In our experience, ∼1% of all patient tumors fell into this category at our institution from 2015-2020 (Methods). Given the high grade, dedifferentiated appearance of these tumors, we reasoned embryonic mapping might provide a new diagnostic approach to determining their origins and could provide a classification. Thus, we applied the D-MLP classifier to 20 such CUP cases, representing the most challenging diagnostic dilemmas seen at our institution.

**Figure 6.**
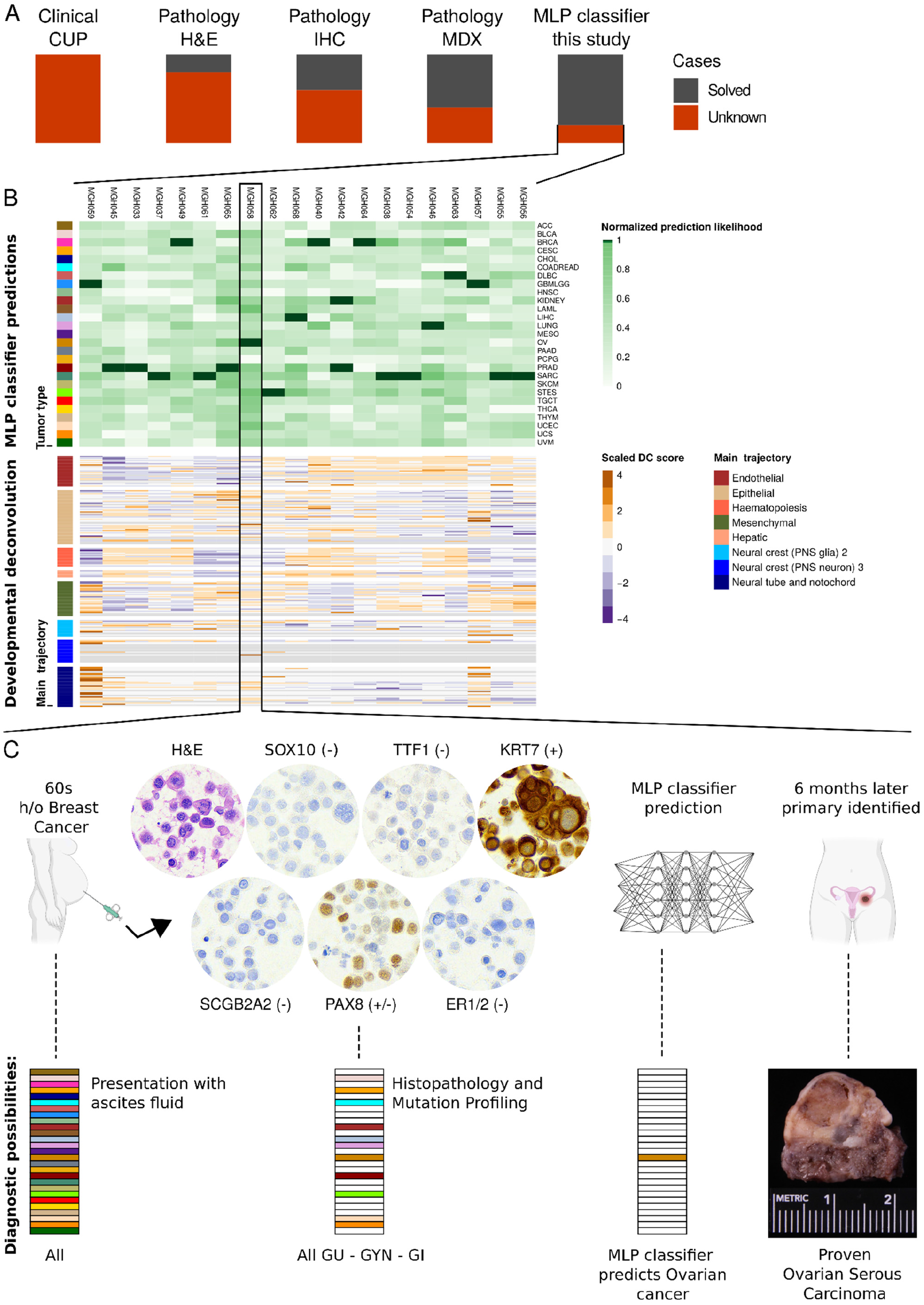
Diagnosis of Cancer of Unknown Primary by a Developmental Multilayer Perceptron D-MLP. **A.** Patients present clinically with CUP. A subset of these cases is solved by hematoxylin and eosin (H&E) examination of tissue. Additional cases are solved by immunohistochemistry stains (IHC) that mark particular tissue types and by molecular techniques (MDX) such as mutation profiling. However, a subset of CUP remains undiagnosed with no primary site assigned after all available techniques. We applied D-MLP to this diagnostically most challenging subset of cases. **B. (Top)** D-MLP classifier predictions for tumor type for 20 CUP cases, shown as classifier likelihood predictions. Note higher confidence predictions are weighted in coloration (dark green). **(Bottom)** The developmental deconvolution profile of each CUP case is shown (developmental component score). Trajectories are arranged top to bottom as they are shown counter-clockwise (Fig. 4 and see Table S4). Colors for tumor type prediction and main developmental trajectory are as in Figures 1–5. **C**. Case MGH058 is highlighted for further consideration. A female patient in her 60’s with a history of breast cancer presented with ascites fluid accumulation. Fluid was drained and assessed by standard cytological/histopathological workup. Stains are shown for H&E (morphology), mammaglobin A (SCG2A2, breast cancer), estrogen receptor (ER1/2, breast cancer), TTF1 (lung), SOX10 (melanoma), Cytokeratin 7 (KRT7, epithelial origin), and PAX8 (broad marker, primarily genitourinary), which ruled out breast cancer but left a broad differential diagnosis encompassing genitourinary (GU), gynecological (GYN), and some gastrointestinal (GI) malignancies. MLP classifier gave a strong prediction of ovarian cancer for this patient’s ascites. 6 months later, after extensive workup, the patient underwent bilateral salpingo-oophorectomy and was found to have a mass (pictured) identified as ovarian serous carcinoma.

The classifier made strong diagnostic predictions for most cases, spread amongst the classification types (Fig. 6B, top). Developmental deconvolution of CUPs revealed that endothelial trajectories contributed strongly to about half of cases; likewise, mesenchymal trajectories contributed strongly to about half of cases though these were not necessarily the same set. Neural tube/notochord trajectories contributed strongly to two cases (MGH059, MGH057), whereas one main trajectory group (neural crest 3) had little contributory information (Fig. 6B, bottom).

If available clinically, this developmental information could have supplemented diagnostic decision-making. For example, case MGH058 was of a female patient in her 60’s with a history of breast cancer who presented with peritoneal ascites. Fluid was drained and examined by cytology, revealing high grade features (nuclear pleiomorphism, mitotic figures) on H&E (Fig. 6C). Immunohistochemistry stains were negative for breast markers (mammaglobin A “SCGB2A2”, estrogen receptor “ER1/2”), lung (TTF1), and melanoma (SOX10), positive for epithelial marker (cytokeratin 7, CK7 or “KRT7”), and variable/weakly positive for genitourinary origin (PAX8) (Fig. 6C). Molecular profiling was positive for variants in RB1 and TP53. This left a very large differential diagnosis with no further tools to narrow it. Analyzing cells from ascites, the D-MLP classifier gave a strong prediction of ovarian cancer for this case (Fig. 6). Six months later, and after extensive additional clinical workup, the patient was found to have a mass proven to be ovarian serous carcinoma. We conclude deep learning classifiers based on developmental deconvolution could serve as a useful adjunct, impacting diagnosis and clinical decision making.

## Discussion

Our analysis systematically compared TCGA tumor samples and MOCA single cell data to construct a developmental map of human tumors. We used this map to deconvolute tumors into developmental components, which in turn allowed us to construct a D-MLP classifier capable of high accuracy tumor type prediction. Together, this constitutes a proof-of-principle for how developmental mapping can be used to aid diagnostic pathology, and for how emerging single cell datasets could impact clinical cancer care as references atlases for diagnostic tools.

Many clinical cases remain diagnostic challenges, and while image-based tools have shown great promise in narrowing differentials, they rely on visual input available to pathologists using current methodologies. Gene expression has the potential to add orthogonal information, but generating models with true predictive power has been difficult, owing to the challenge in extracting the most relevant information. Our approach used developmental trajectories to dimensionally reduce gene expression data. Projecting tumor data onto axes of reduced dimensionality defined through developmental programs, instead of defined through gene expression, markedly increased accuracy of classification. Another benefit of a developmental approach is that this focus can reveal new tumor biology, such as heterogeneity within or between tumor types. However, while powerful, this approach has drawbacks, as not all information germane to tumor classification may be developmentally related. An additional limitation is that the current study primarily utilized TCGA data, which are of relatively high tumor purity compared to clinical samples. In fact, admixed tumor-normal samples can yield diagnostic information picked up in bulk gene expression, such as tumor types more likely to be vascularized or with particular admixed histology, and this can aid in the classification of CUPs as they may show the same admixed tissue types, helping signal a particular diagnosis. Additionally, the current reference dataset was from a different species (*Mus musculus*), as this was the largest single cell developmental reference atlas available at the time of study and therefore captured the greatest amount of developmental heterogeneity. While mice have proven useful models for human cancer, it will be of great interest to construct new classifiers using human single cell datasets, such as those that may emerge from the Human Cell Atlas (45). The approach described here should easily generalize to such datasets as they become available.

The approach in the present study focused on broad categories of malignancies. Yet, many current diagnostic dilemmas in pathology focus on distinguishing different entities within one tissue type rather than distinguishing different tissue types from one another. New classifiers, trained on extensive datasets within one tissue type, may be able to distinguish these entities on the basis of their development. Our comparisons suggested underlying differences in developmental components between tumor and normal tissue. Classifiers focused on distinguishing benign entities from malignant ones would be of great use in pathology, especially in histopathological ‘gray zone’ cases, as clinical management decisions often turn on whether an entity is classified as benign or malignant. In TCGA, not all patients have normal samples matched to tumor, and thus TCGA may not adequately represent unaffected regions of all patients. As larger cohorts of normal samples become available these will boost diagnostic accuracy when used as reference datasets for classifiers that focus on distinguishing benign from malignant entities.

One challenge in building deep machine learning classifiers for CUPs, especially for the most challenging cases, is that by definition no current gold standard exists against which to compare predictions. Perhaps a combination of models, such as those that analyze both imaging data and molecular features, will prove to be most useful in achieving precision cancer care. Ultimately, prospective studies will need to be done to demonstrate the benefit of machine learning approaches. The results presented here give a developmental map of human tumors, and suggest a new tool for decreasing diagnostic uncertainty in pathology with implications for diagnostic classification of cancer.

## Supporting information

Table S1: TCGA cohort samples used in this study given by TCGA sample code

Table S2: Summary statistics for correlation coefficients shown in Figure 2 by tumor type

Table S3: Pan-cancer cohorts used for analyses in Figure 3

Table S4: Developmental Component Scores by sample, and order of developmental components on figures

Table S5: Full cohort of samples from multiple studies used for construction of D-MLP, and list of samples held out to test classifier performance

Table S6: Gene name conversions between mouse and human samples along with TCGA gene IDs

Table S7: Conversion of non-TCGA study tumor types to codes used in D-MLP classification

Table S8: Hyperparameter optimization for D-MLP and genes used for benchmark classifiers

## Data Availability

Code is available at https://github.com/emoiso/DevTum. Original data are available on TCGA and MOCA websites and as described further in the manuscript. All other data produced are available upon request to the authors.

## Acknowledgements

We thank Kelli Burke for administrative assistance. We thank Phillip A. Sharp, Jacqueline A. Lees, Amanda J. Whipple, and members of their laboratories for helpful discussions. This work was supported by a Charles W. and Jennifer C. Johnson Clinical Investigator Award (SG), NCI K08-CA237856 (SG), NCI R37-CA225655 (JKL), and NCI P30-CA14051 (Koch Institute core). EM acknowledges support from a Ludwig Fellowship (Koch Institute). **Human Studies** This work was performed under Mass General Brigham IRB #2014P000940 and MIT COUHES E-2066. **Author contributions**: EM and SG designed the study. AH, SM, and SL developed FFPE sequencing techniques. AF and HM selected and provided samples. JL and SG interpreted cases. EM developed the models and performed the analysis. SG supervised the research. EM and SG wrote the manuscript with input and approval from all authors.

## Data and code availability

Described in materials and methods below.

---

**Supplementary Figure 1:**
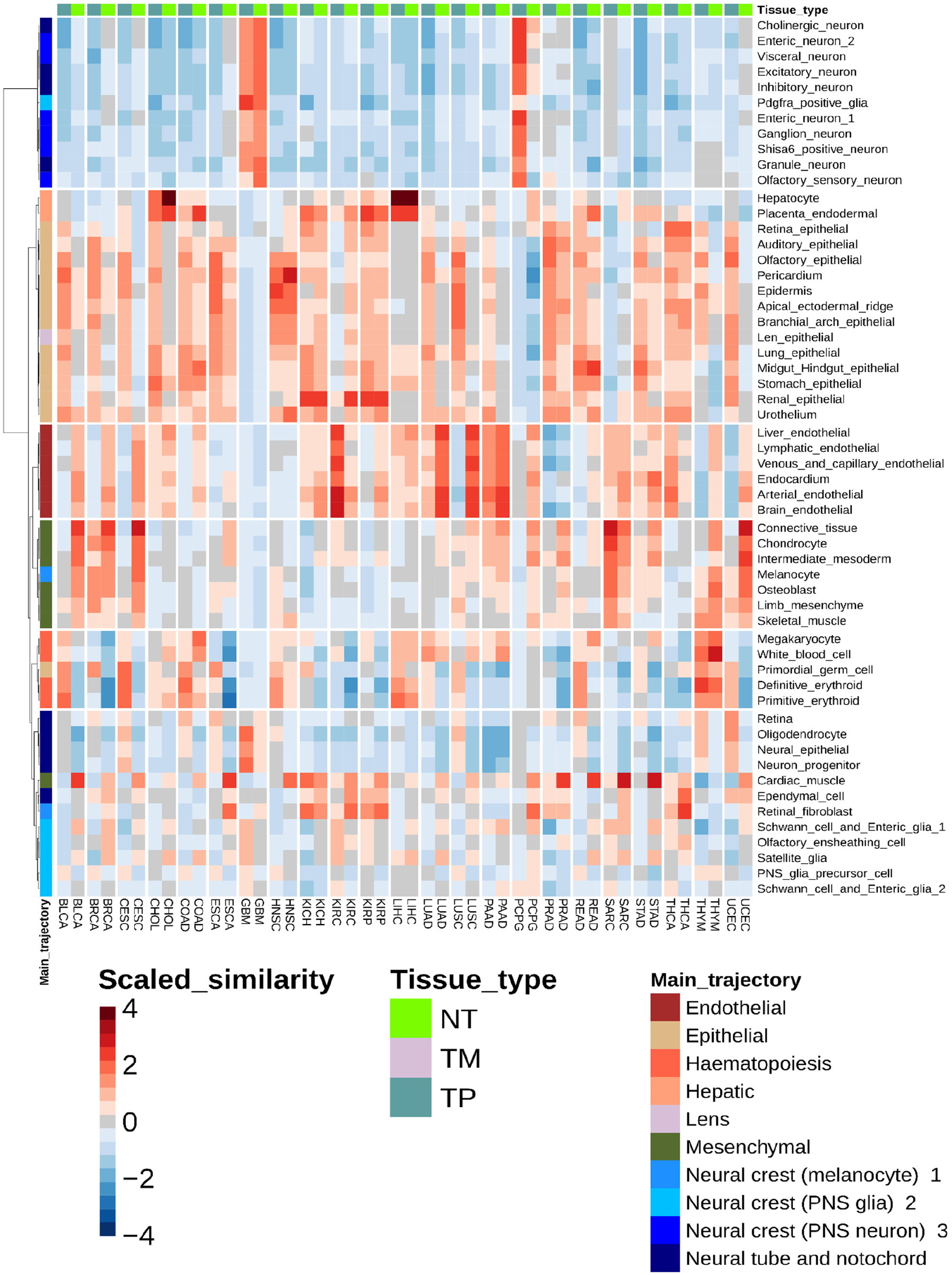
Tumor-normal tissue type similarity with developmental trajectories. Shown is a heatmap equivalent to main Fig. 1C, arranged by placing each primary tumor adjacent to the equivalent normal tissue for all patient sample types for which this data was available in TCGA. Main and sub-trajectories and scaled similarity scores are as in Fig. 1C.

**Supplementary Figure 2:**
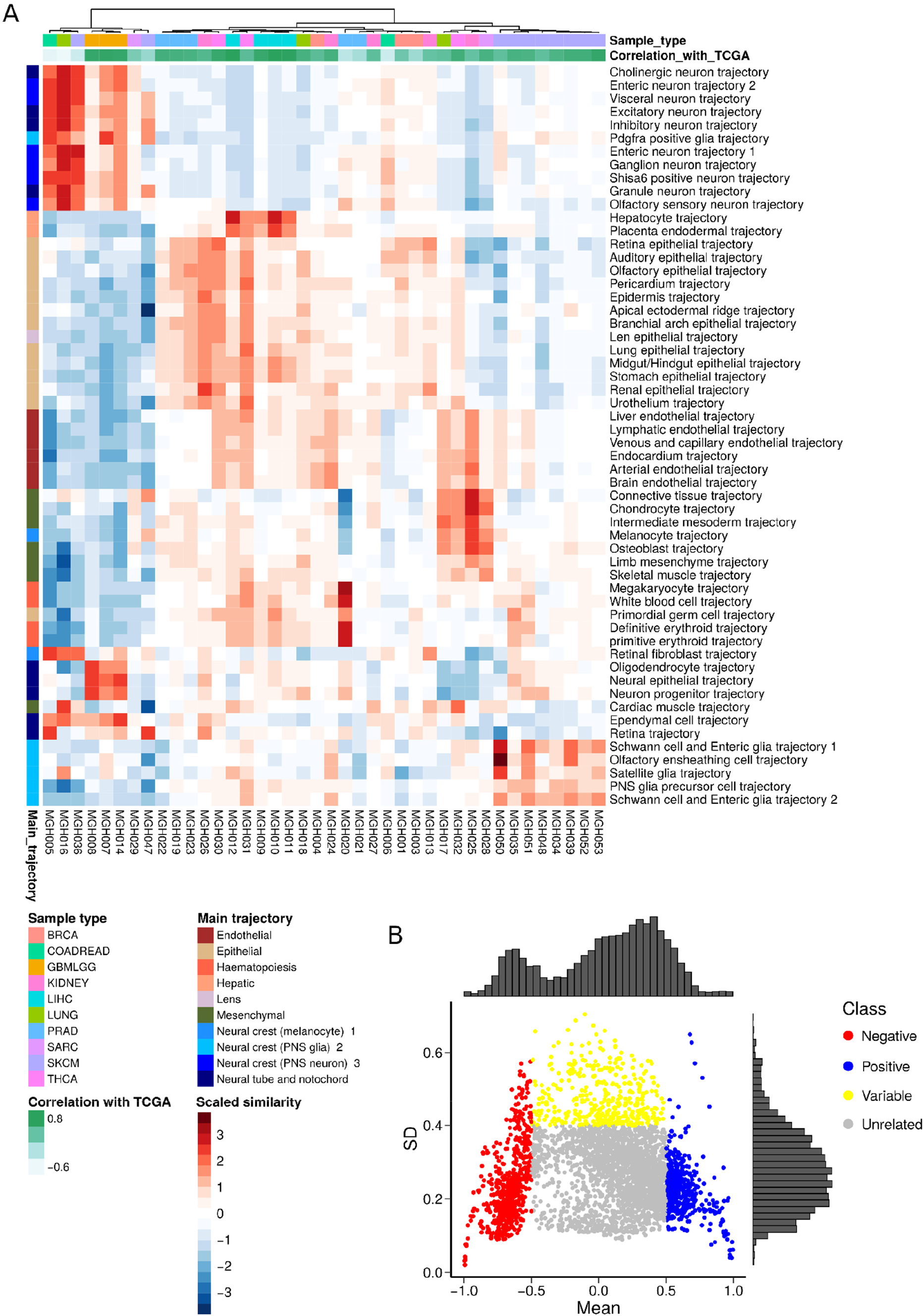
Validation of TCGA and MOCA correlations using clinical samples and further definition of relationships between trajectories and tumors. **A**. We sequenced the transcriptome of 40 tumors with known classification using nucleic acid (RNA) isolated from FFPE tissue. We performed scaled correlation analysis for this cohort against MOCA trajectories identical to the procedure for TCGA gene expression. Shown are the known sample type (histopathological diagnosis, top row) and the spearman correlation coefficient (second row) for the rank order of trajectory correlation for each MGH FFPE sample (columns) against the equivalent TCGA primary tumor type (Fig. 2C). The average spearman ρ across all multiple hypothesis corrected correlations is 0.69. See Methods for further details. Note, two samples (far left) appear anti-correlated when comparing FFPE correlated trajectories to TCGA correlated trajectories; removal of these two from the analysis raises the ρ to 0.76. **B.** The correlation coefficients between all cells in a MOCA developmental sub-trajectory and all TCGA tumor types were plotted as similarity scores (see Fig. 1B, right). Then, the mean and standard deviation of the similarity score distribution was calculated and plotted above. Thus, each dot on the graph represents the relationship between one sub-trajectory and one tumor type. Relationships that had low mean similarity scores and low deviation were defined as unrelated/uninformative. Relationships characterized by high similarity scores (such as inhibitory neuron trajectories with LGG) were defined as positive (blue), relationships with strong negative mean similarity scores (such as inhibitory neuron trajectories with LIHC) were defined as negative (red), and relationships with low mean similarity scores but high variance across cells were defined as variable (yellow). Note the latter means that within the trajectory, there are cells with strong positive correlations to tumor type and strong negative correlations to tumor type. Relative density histograms are shown on top for mean values and at right for standard deviation values.

**Supplementary Figure 3:**
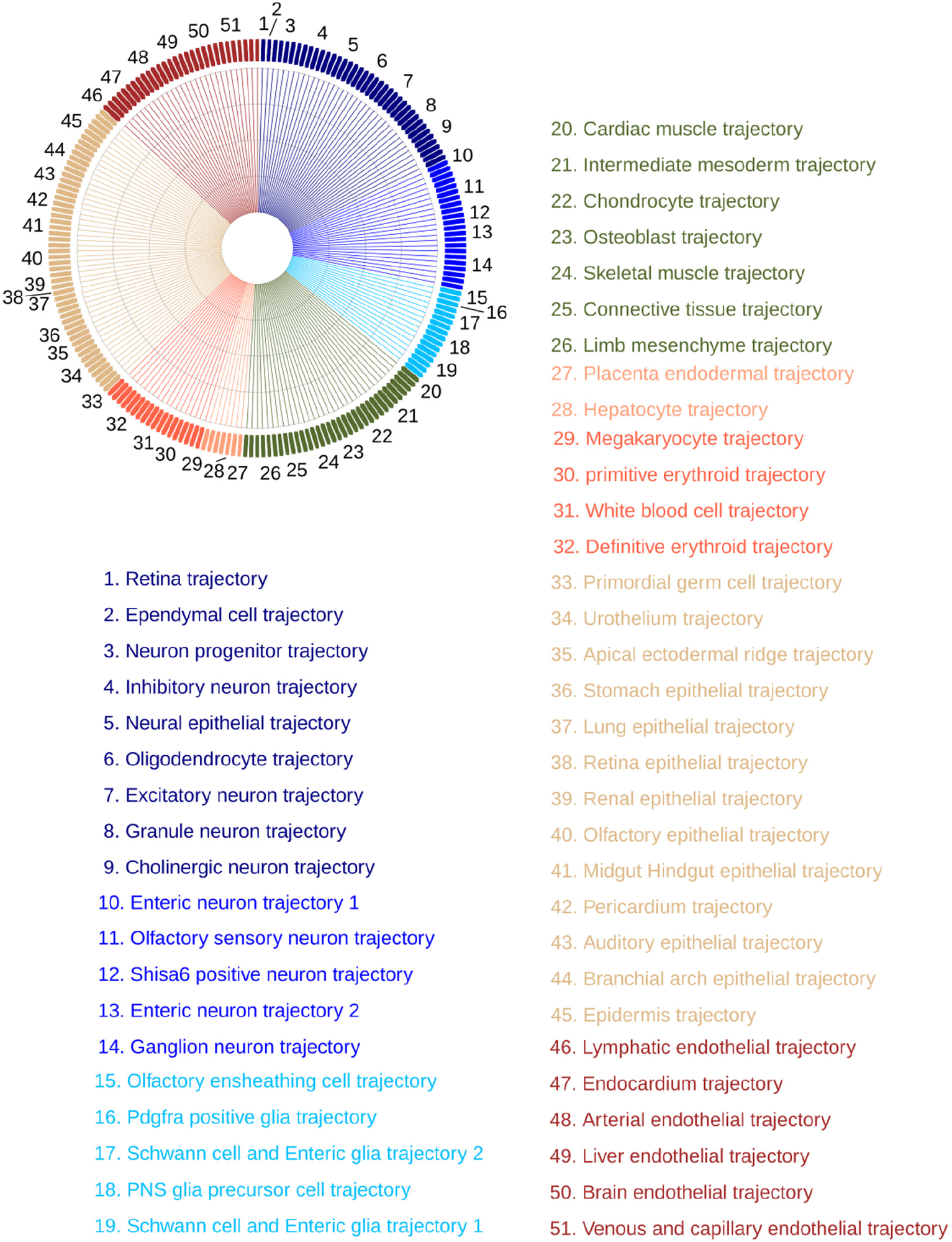
Sub-trajectories location on radar plots. Above, order and location of sub-trajectories on radar plots (Fig. 4) and the order of sub-trajectories shown in Fig. 6B is indicated. For each sub-trajectory, deconvolution scores are calculated at each timepoint. Note that while 56 sub-trajectories appear in the MOCA dataset not all of these contributed to deconvolution scores of TCGA samples; those that did not contribute (no cells in the trajectory were amongst the highest correlated to any sample) were omitted from the plot for clarity. A total of 214 contributory developmental components across timepoints of 51 sub-trajectories were used, see Table S4 for a complete list.

**Supplementary Figure 4:**
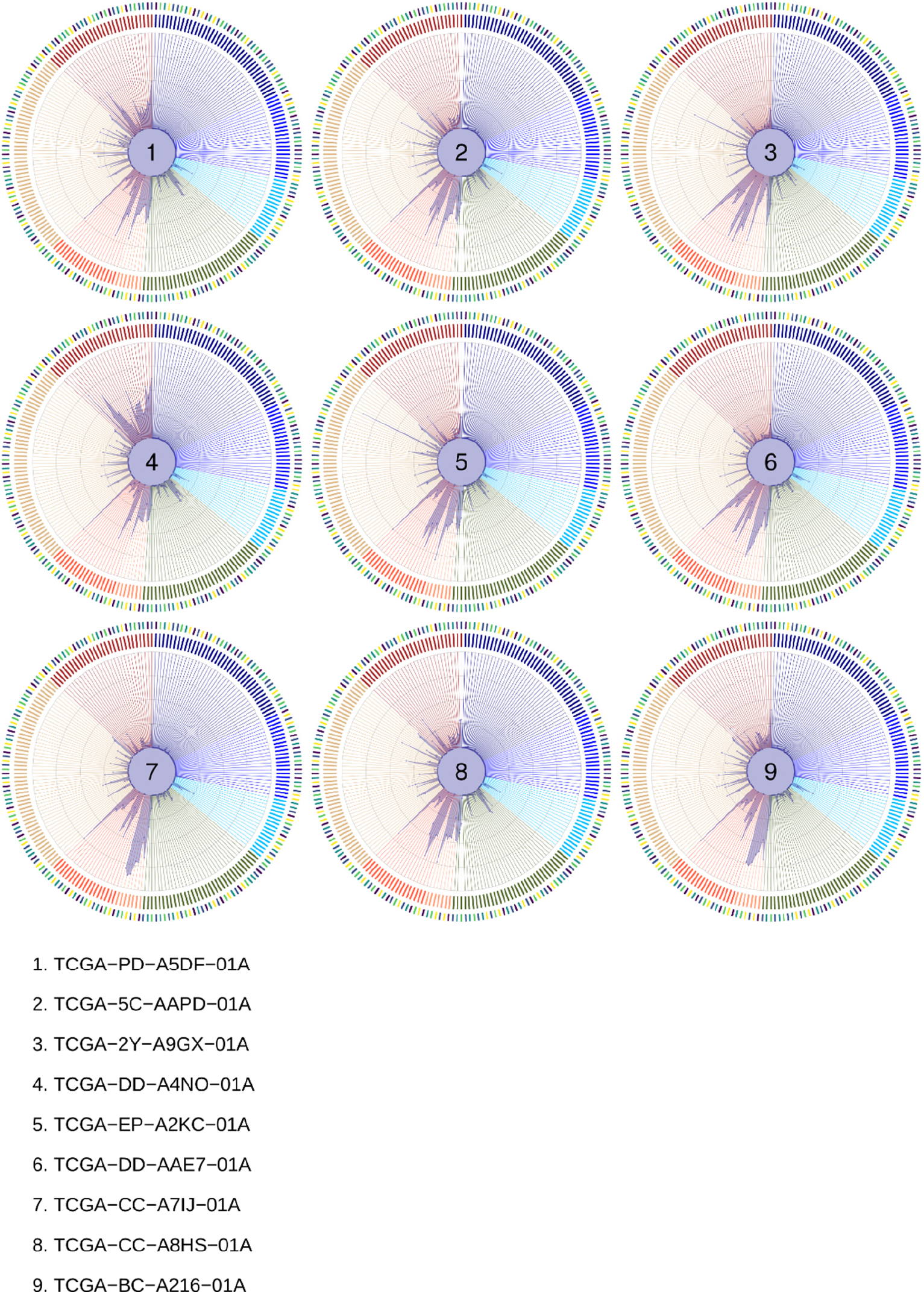
Developmental deconvolution radar plots for nine randomly chosen hepatocellular carcinoma samples. Note that hepatic trajectories (peach, 7 o’clock) show strong signal in all samples whereas endothelial trajectories (maroon, 11 o’clock) show variable signal between samples. Neuronal and neural crest trajectories (blue, 12 o’clock to 4 o’clock) show low signal in all samples.

**Supplementary Figure 5:**
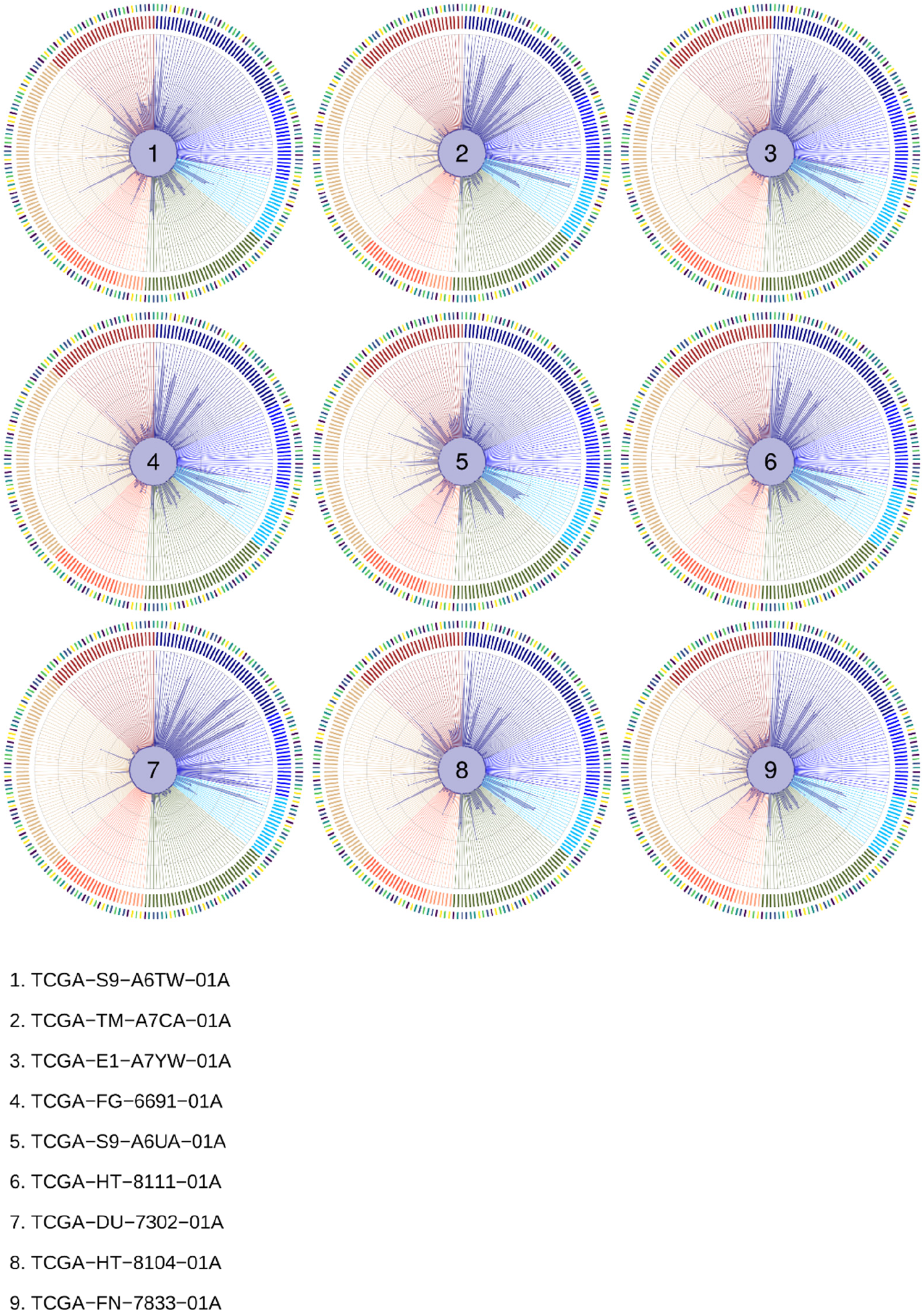
Developmental deconvolution radar plots for nine randomly chosen low grade glioma samples. Note that hepatic trajectories (peach, 7 o’clock) show no signal in all samples and endothelial trajectories (maroon, 11 o’clock) show very little signal in all samples. In contrast, neuronal and neural crest trajectories (blue, 12 o’clock to 4 o’clock) show high signal in all samples, and the identity of which trajectories contribute to each sample varies significantly.

**Supplementary Figure 6:**
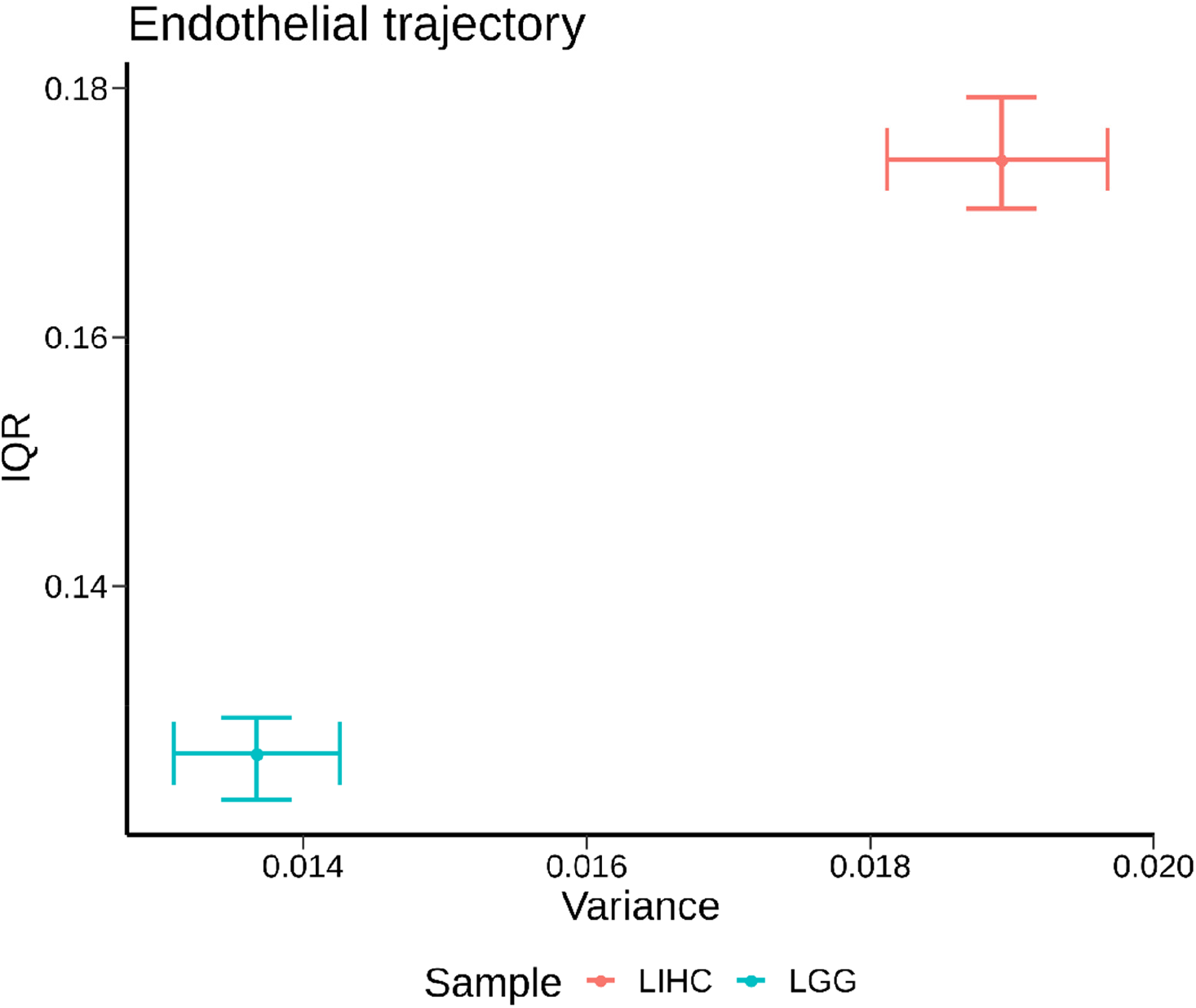
Summary of variance across hepatocellular carcinoma and low-grade glioma samples for endothelial trajectory developmental deconvolution scores. Plotted are the variance across samples for endothelial DC scores versus the inter-quartile range (IQR, difference between the median of the upper half of the data minus the median of the lower half of the data). Dots indicate the mean of each value and error bars the 95% confidence interval (2.5^th^-97.5^th^ percentiles across samples). To obtain a single endothelial DC score for each sample, the DC scores across all sub-trajectories at all timepoints that fell into main category endothelial were first average for each sample. Note the axes do not start at 0 (in order to aid visualization of the spread across samples).

**Supplementary Figure 7:**
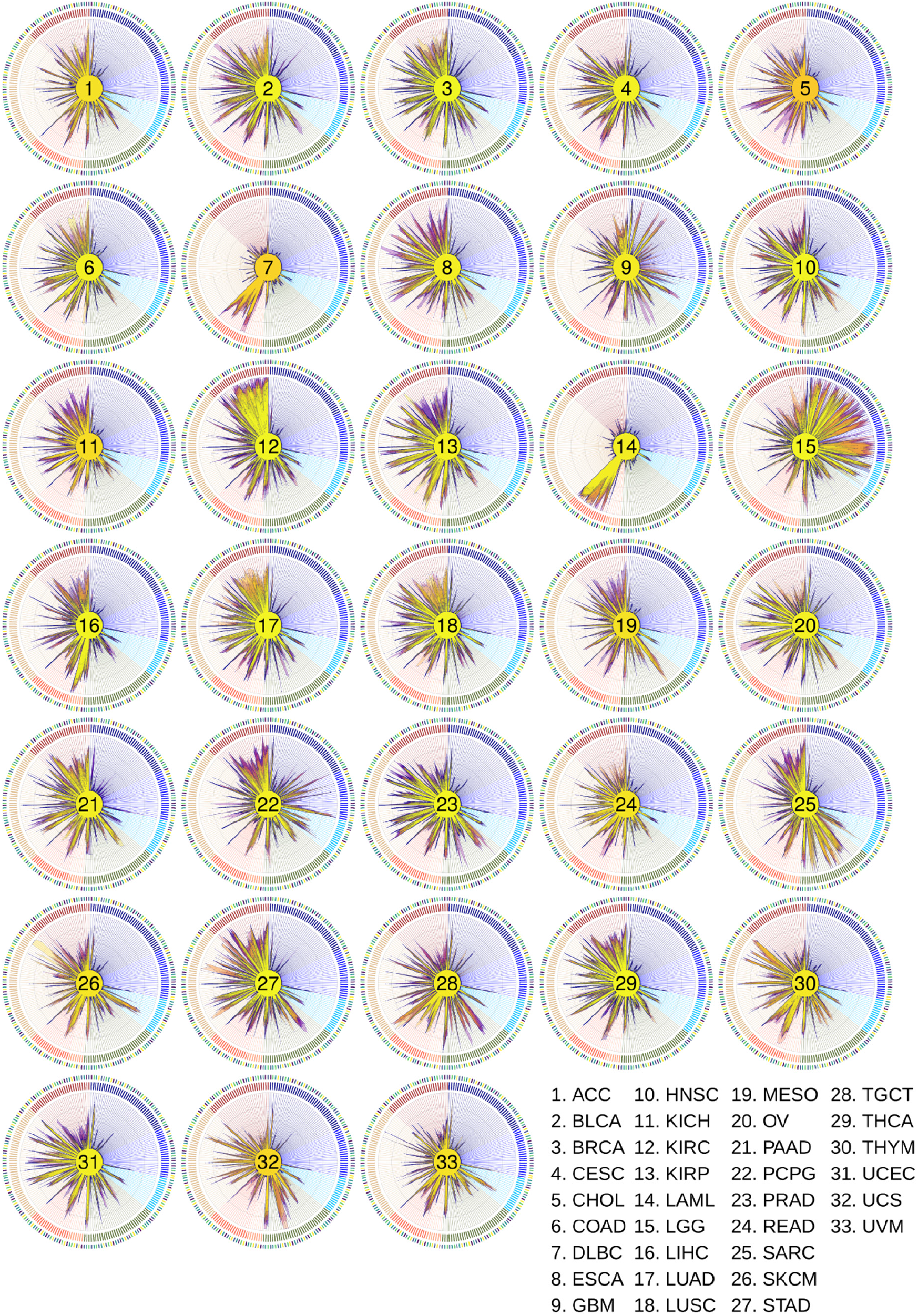
Summary radar plots for all TCGA tumor types. Shown are radar plots summarizing the deconvolution profiles of all samples for each TCGA tumor type.

**Supplementary Figure 8:**
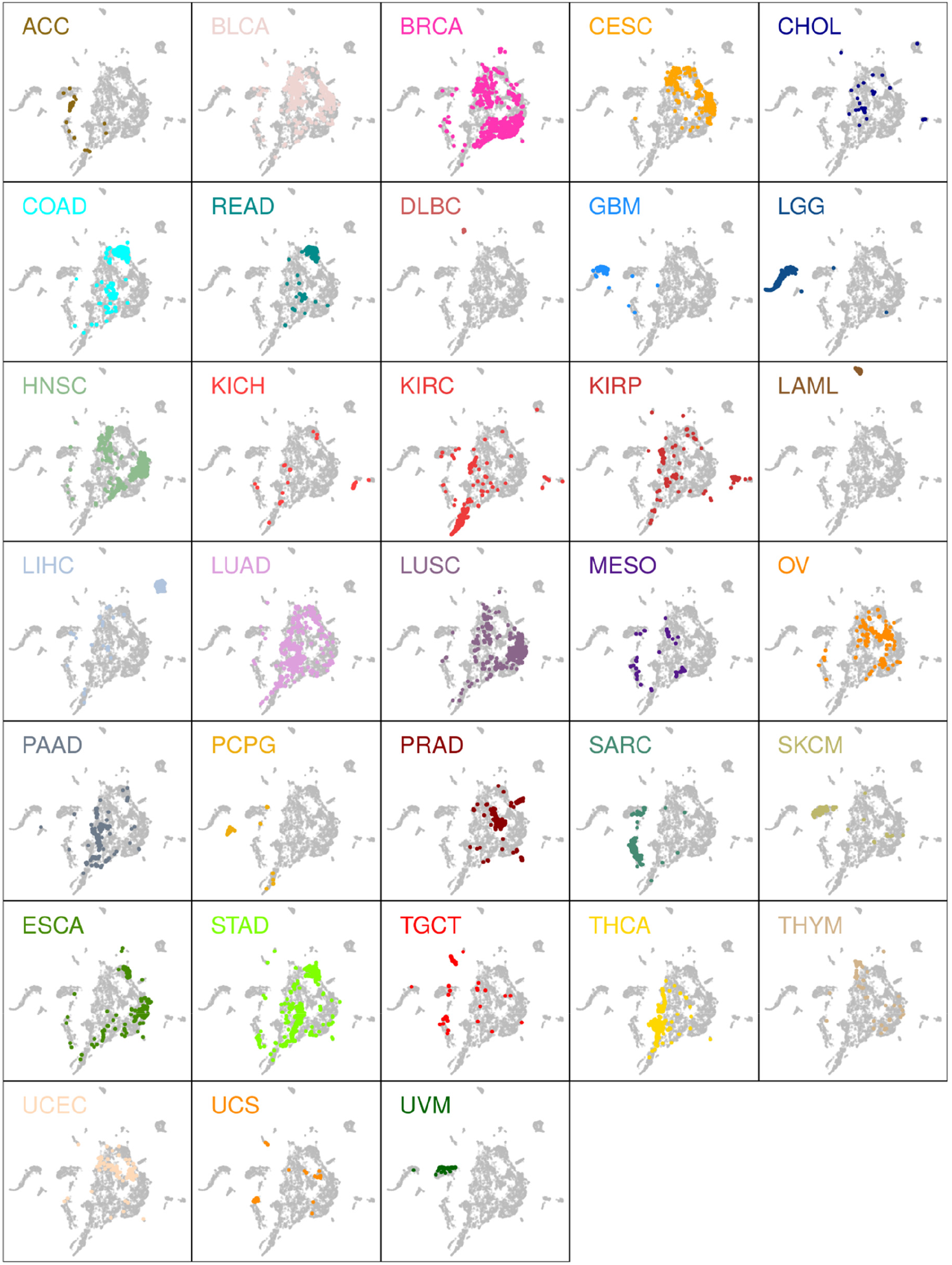
UMAP representation of developmental deconvolution scores highlighted by tumor type. Highlighted in each panel are the samples belonging to each of the 33 TCGA sample types. Primary tumor, metastasis, and normal tissue for each sample type are highlighted. Position of samples is equivalent to Fig. 4F-G.

**Supplementary Figure 9:**
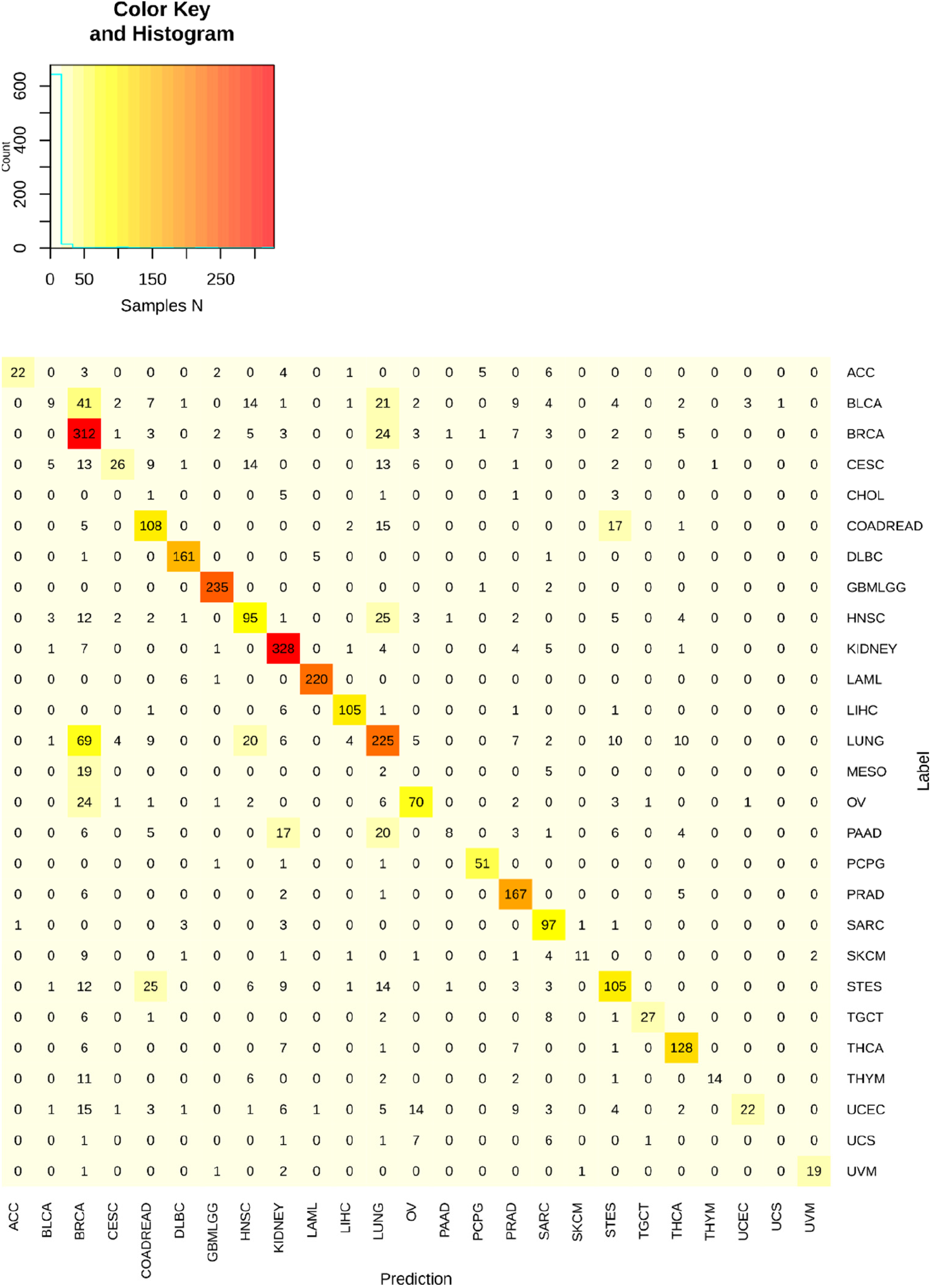
Confusion matrix for MLP performance. Shown above are all top1 prediction results for the MLP classifier. The y-axis represents the sample tumor type as assigned by the study from which it was obtained, and x-axis the tumor type labels for the prediction of the classifier. On diagonal results represent true positives. Note the matrix is not perfectly square as CHOL and MESO are omitted from columns as the classifier never predicted these outcomes (0 columns).

**Supplementary Figure 10:**
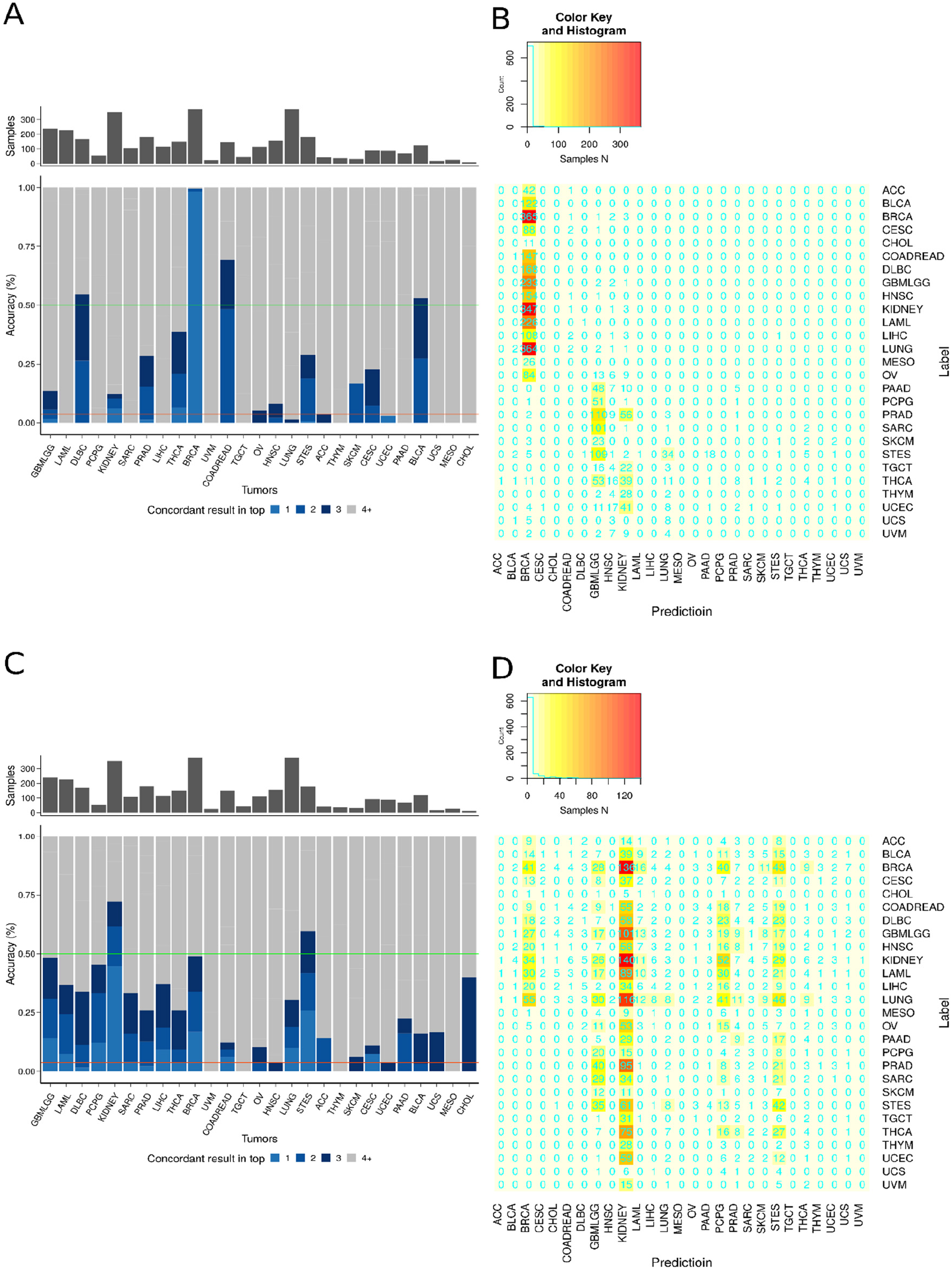
Performance of ‘Highly Variable Genes’ and ‘Clinical Oncopanel Gene’ classifiers. **A + B**. The accuracy (A) and confusion matrix (B) of the ‘highly variable genes’ classifier are shown for each tumor type. **C + D**. The accuracy (C) and confusion matrix (D) of the ‘clinical oncogenes panel’ classifier are shown for each tumor type. Ordering of tumor types are identical to Figs. 5C and S9 to facilitate comparison. For Highly Variable Genes classifier, top1 accuracy is 10.6% and top3 22%. For Clinical Oncopanel Genes, top1 accuracy is 7.9% and top3 accuracy is 21%.

**Supplementary Figure 11:**
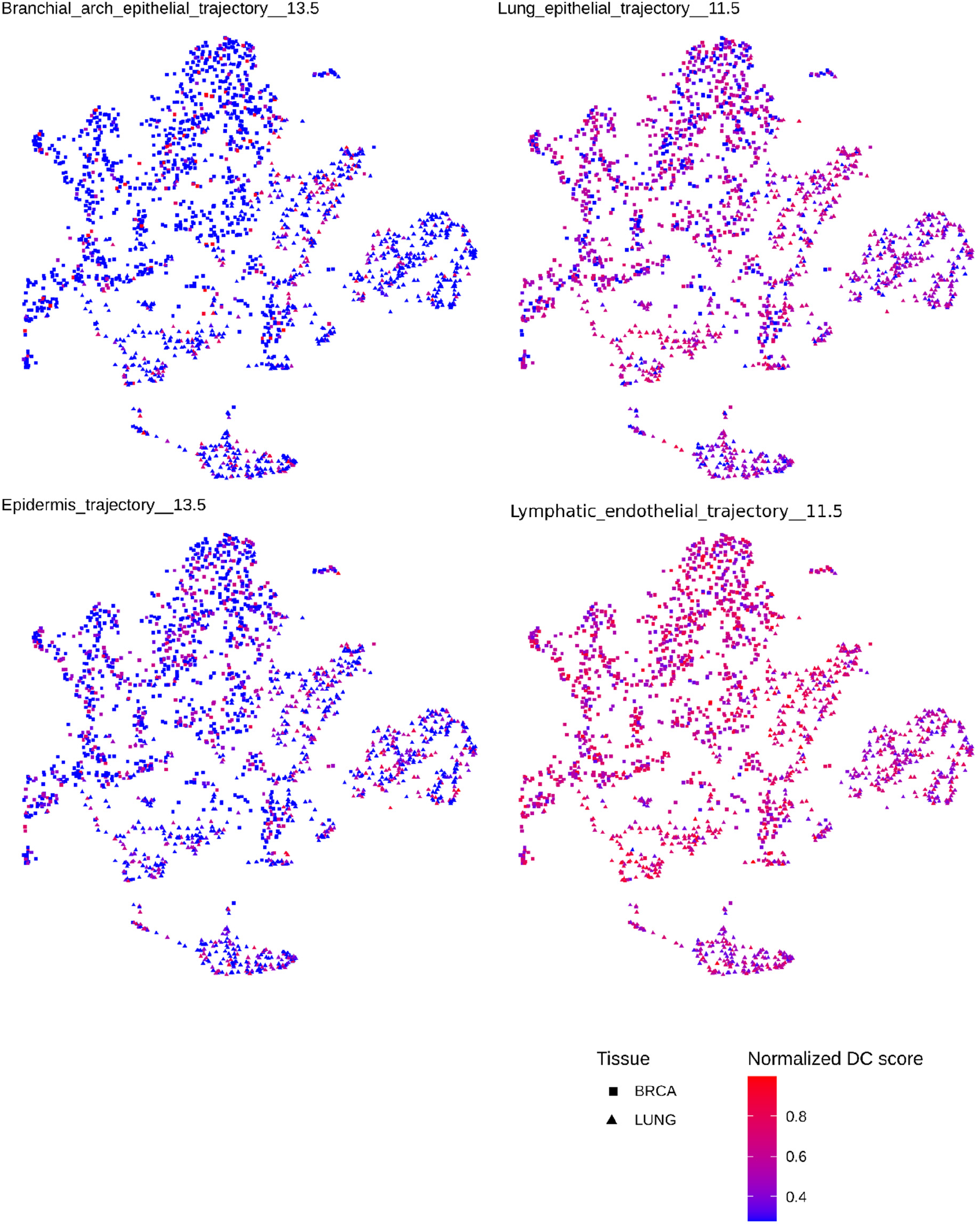
Heterogeneity in Breast and Lung Tumors for Particular Developmental Trajectories. Breast (squares) and lung (triangles) cancer samples from the cohort are shown by UMAP performed across all of their developmental component scores. Samples are then colored separately by their DC scores for the 4 indicated developmental trajectories. Heterogeneous patterns for all 4 components across both breast and lung cancer samples is observed, suggesting shared developmental programs in these two tumor types.

**Supplementary Figure 12:**
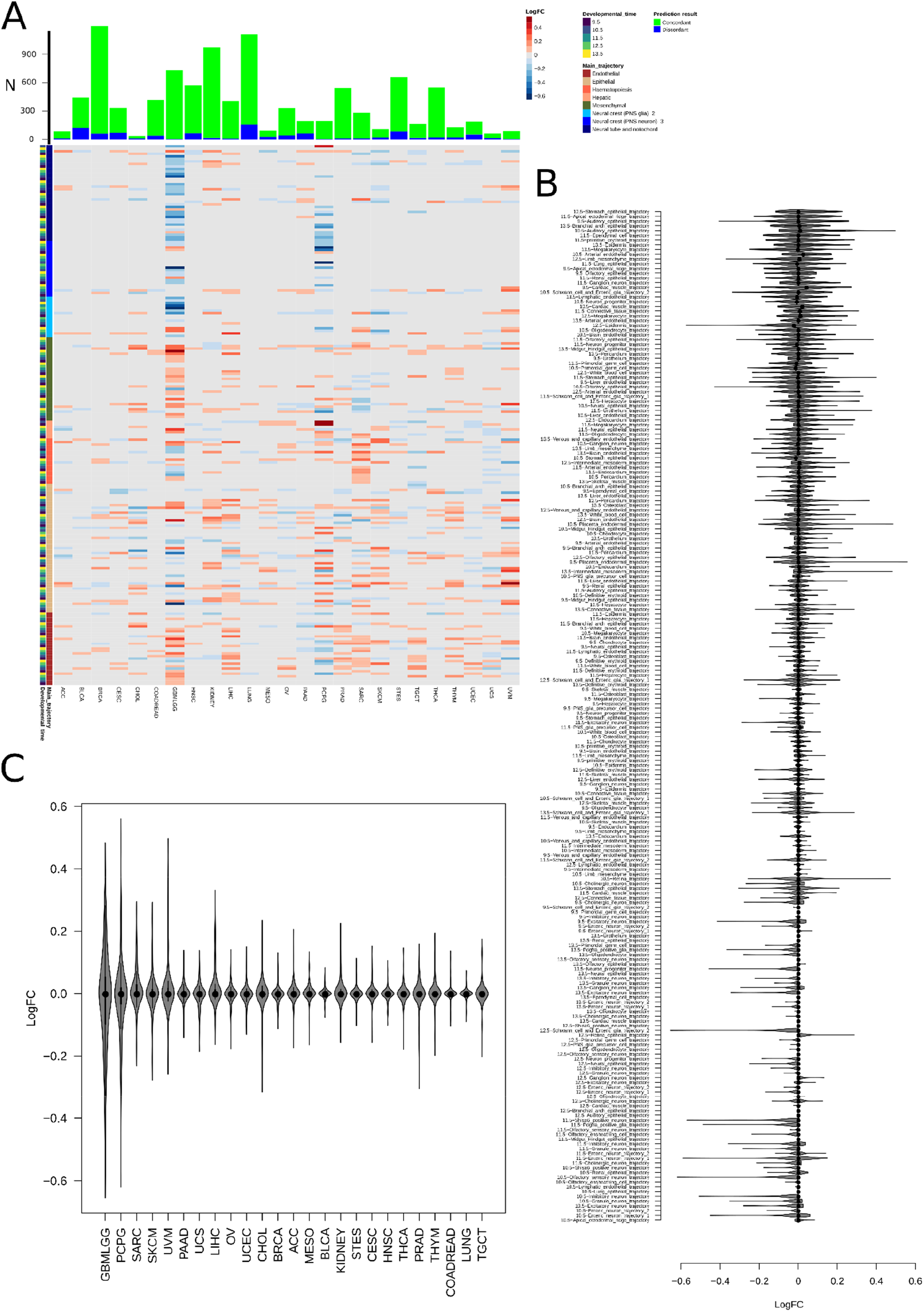
Analysis of discordant MLP classifier predictions. **A**. Log_10_(fold change) for the contribution of each developmental trajectory (214 total, rows) in concordantly classified samples to discordantly classified samples for each tumor type. The main trajectory and developmental time window for each trajectory is shown at left. The number of concordantly and discordantly classified samples inputted into the fold change calculation is displayed above. Log(FC) values plotted are for adjusted p-values < 0.05 (using linear model estimation, R limma package), all fold changes with adjusted p-value > 0.05 are set to 0. Trajectories are ordered as given in Table S4. **B**. All 214 contributory trajectories are listed and the log_10_fold change between concordantly classified samples and discordantly classified samples is plotted. Each data point represents a single tumor type (analyzed as a log ratio of signal for the trajectory in concordantly classified tumors to discordantly classified tumors). **C**. All classified tumor types are listed and the log_10_fold change between concordantly classified samples and discordantly classified samples is plotted. Each data point represents a single trajectory (analyzed as a log ratio of signal for the trajectory in concordantly classified tumors to discordantly classified tumors). Violin plots represent the median (line) with quartiles (edges: 25^th^-75^th^ percentiles, whiskers: 25^th^ percentile minus IQR to 75^th^ percentile plus IQR).

## Materials and Methods

### Data gathering and Sample Cohorts

#### MOCA

The expression profile and meta information of cells analyzed in (MOCA (Cao et al., 2019)) RNA sequencing [gene_count_cleaned.RDS] and annotation [cell_annotation.csv] data were manually downloaded from: http://atlas.gs.washington.edu/mouse-rna/downloads. For this study, we used expression data of the 1,331,984 high quality cells as defined in the MOCA study. Briefly the cells were filtered as follows: less than 400 detected mRNA molecules, detected doublets, and all cells from doublet derived sub-clusters were removed from analysis.

#### TCGA

The coding gene expression profile [RNAseqV2_RSEM_genes_normalized data_Level_3] and clinical information [Merge_Clinical.Level_1.2016012800.0.0] of TCGA samples (release 2016_02_28) were systematically downloaded using *firehose_get* v 0.4.1 tool, downloaded from the Broad TCGA GDAC site (https://confluence.broadinstitute.org/display/GDAC/Home). The clinical information files were further filtered to retrieve histological grade information. In cases where multiple samples from the same patient were taken, these were classified as primary tumors, normal tissue, or metastatic samples respectively as appropriate and analyzed separately. No cases were present with multiple normal, primary, or metastatic samples from the same tumor in the cohort.

#### NON TCGA Sample Cohorts

The “NON TCGA” cohort refers to samples contained in various cancer studies, for which gene expression profiles and clinical information can be retrieved from the Genomic Dara Commons (GDC) Data Portal (https://portal.gdc.cancer.gov/). The full list of samples and relative studies used in this work and labelled as “NON TCGA” is given in Supplementary Table 6, as well as their conversion to 27 TCGA diagnostic categories for purposes of classification. In brief, TCGA categories COAD and READ were merged into COADREAD, GBM and LGG into GBMLGG, KICH KIRC and KIRP into KIPAN, LUAD and LUSC into LUNG, and ESCA and STAD into STES. Merging of TCGA categories facilitated incorporation of a maximum of non-TCGA samples, as these used slightly different diagnostic categories. A list of TCGA study abbreviations is given at (https://gdc.cancer.gov/resources-tcga-users/tcga-code-tables/tcga-study-abbreviations).

#### MGH Sample Cohort

Samples from formalin fixed paraffin embedded tissue were chosen from cases seen in the Center for Integrated Diagnostics in the Department of Pathology at Massachusetts General Hospital either with known diagnosis (40 cases) or as cancers of unknown primary (20 cases). Total nucleic acid was isolated from six scraped blank slides using clinically validated protocols. At MGH over the period 2015-2020, 29,669 tumors were seen as reported by our institution to mandatory reporting agencies. Of these, 208 tumors (=0.7%) never received a primary site through their final diagnosis as estimated by case coding. 20 of these cases were retrievable with extracted nucleic acid for further testing by the developmental deconvolution classifier and their analysis is the subject of Fig. 6.

### RNA extraction from FFPE Clinical Samples

Libraries were prepared using a modified version of the Takara SMARTer Stranded Total RNA-Seq Kit - Pico Input Mammalian kit. In brief 100ng of RNA at 10ng/ul was sonicated using RL230 Covaris sonicator (Covaris Inc) and resulting material was confirmed using a Fragment Analyzer (Agilent). 10ng of each sonicated sample was prepared using the pico input kit as for FFPE samples using a 1:8 volume reduction on the STP MosquitoHV. Final libraries were validated by Fragment Analyzer and qPCR prior to sequencing on a NovaSeq6000 S4 with 150nt paired-end reads.

### FFPE Clinical Samples derived RNA-Sequencing analysis

Reads obtained from the sequencing step previously described were first aligned to the genome and subsequently gene counts calculated as follows. Alignment was performed against the Homo Sapiens primary assembly v35_GRCh38.p13 genome and relative annotation GTF file (v35), downloaded from GENCODE website (https://www.gencodegenes.org/) and further processed to be compatible with STAR v2.7.1a (46) alignment tools and RSEM v1.3.1 (47) for gene expression analysis. The details of the commands and parameters used to generate the gene expression matrix from fastq are contained in mgh_sequencing.sh file. In brief, reads were processed as follows. The genome fasta and annotation has been processed to be compatible with RSEM for expression calculation with the *rsem-prepare-reference command* with the following options: -p 30, --star; and a STAR compatible genome files as follows: STAR -- runMode genomeGenerate --runThreadN 30 --genomeFastaFiles GENCODE/v35_GRCh38.p13/GRCh38.p13.genome.fa –sjdbGTFfile GENCODE/v35_GRCh38.p13/gencode.v35.annotation.gtf --sjdbOverhang 149. Then, a first pass STAR alignment was performed to achieve accurate information on gene exon-intron boundaries, as follows: STAR --runThreadN 16 --outFilterType BySJout --outFilterMultimapNmax 20 --alignSJoverhangMin 8 -- alignSJDBoverhangMin 1 --outFilterMismatchNmax 999 --outFilterMismatchNoverLmax 0.04 -- alignIntronMin 20 --alignIntronMax 1000000 --alignMatesGapMax 1000000 –readFilesIn $[file]_1_sequence.fastq $[file]_2_sequence.fastq. This led to the generation of the first alignment files that were subsequently used for the generation of a second reference genome as follows: STAR --runMode genomeGenerate --runThreadN 30 --genomeFastaFiles GENCODE/v35_GRCh38.p13/GRCh38.p13.genome.fa --sjdbGTFfile GENCODE/v35_GRCh38.p13/gencode.v35.annotation.gtf --sjdbFileChrStartEnd *_SJ.out.tab -- limitSjdbInsertNsj 3000000 --sjdbOverhang 149. The second alignment was performed to generate an output compatible with the *rsem-calculated-expression* mode as follows: STAR --outSAMunmapped Within - -outFilterType BySJout --outSAMattributes NH HI AS NM MD --outFilterMultimapNmax 20 -- outFilterMismatchNmax 999 --outFilterMismatchNoverLmax 0.04 --alignIntronMin 20 --alignIntronMax 1000000 --alignMatesGapMax 1000000 --alignSJoverhangMin 8 --alignSJDBoverhangMin 1 --sjdbScore 1 --runThreadN 16 --genomeLoad NoSharedMemory --outSAMtype BAM Unsorted --quantMode TranscriptomeSAM --outSAMheaderHD \@HD VN:1.4 SO:unsorted --outFilterScoreMinOverLread 0.3 -- outFilterMatchNminOverLread 0.3. To generate a gene expression matrix (given in the /Data folder, see below) the following command was used: rsem-calculate-expression -p 16 --bam --paired-end --no-bam-output --forward-prob 0.5 --seed 12345. Gene expression, generated by rsem and measured in transcripts per million (TPM) at the gene level, were then used to evaluate the similarity with MOCA cells as previously described.

### Data preparation

The conversion from Mouse Ensembl id (MOCA) to Human gene symbol/Entrez id (TCGA) was achieved using BioMart (https://www.ensembl.org/biomart/martview/) Ensembl v95. Human gene symbol/Entrez id (TCGA) to Ensembl id (NON-TCGA) mapping was achieved using *org.Hs.egENSEMBL2EG* from org.Hs.eg.db (v3.8.2) Bioconductor (v3.9) (https://bioconductor.org/) package. The intersection of these two sources of information was performed using the Human gene symbol/Entrez id shared identifier. This process generated a list of translated names, given in Supplementary Table 7. This list was then used as a dictionary for gene names and mouse-human ortholog conversion. This identified 15,929 unique human genes that were used in this study. In the case of multiple mouse gene names mapping to the same human gene name, the average expression levels were calculated across occurrences.

### Similarity score calculations

The similarity between MOCA cells and TCGA, NON-TCGA and MGH samples was calculated using a Spearman Correlation coefficient [1], by means of *cor* function in R (see code *correlation.R* for more details), on the expression profile of all shared genes identified as described above for each TCGA sample and MOCA cell:

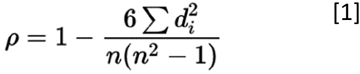

Spearman correlation coefficient, rho ρ, formula [1], *d* is the difference between the two ranks of each observation and *n* is the number of observations. This non-parametric method was chosen as it is more robust to outliers, is unaffected by the normalization method, and allowed a rank-based method less prone to artifacts from single cell transcript dropout. After calculation of each raw correlation score comparing each cohort tumor to each single cell, correlation coefficients were averaged and normalized as follows to generate Figures 2A, 2B and 2C:

I. let ρ_i be a vector of length 1331984, containing the correlation coefficient between human sample human (i) and 1331984 mouse cells belongings to 56 developmental trajectories.
II. ρ_i is mean (mu) centered and standard deviation (sigma) scaled, to obtained a normalized vector for each human sample.
III. Each normalized vector belonging to the same sample type (e.g. all primary breast, all normal breast, all metastatic melanoma etc..) were averaged together, creating a vector of trajectories per tissue and sample type (62 in total).
IV. The scores of each cell from step III were mean centered and sigma scaled across the 62 different tissue and sample type.
V. The cell scores of each trajectory were averaged together reduce the length of the 62 vectors of step IV from 1331984 to 56. This returned the Z-score of the correlation coefficients, also referred to as scaled similarity score in figures 2A, 2B, and 2C. See also code data_recap.R and overall_Z_scores.R

### Deconvolution by developmental components (DCs)

In order to perform developmental component deconvolution, for each tumor sample the 1,331 (∼.1%) most strongly correlated cells were selected. In brief, the cells were ranked in reverse order based on correlation coefficient such as to assign the lowest rank number to the most correlated cells and the highest rank number to the least correlated one (the most correlated cells, ranked first, was attributed the score of 1,331. Conversely, the least correlated of the 1,331 cells, was attributed the score of 1). The ranks were then summed for all the cells with the same developmental identity (i.e. belonging to the same sub-trajectory and developmental time of origin), creating the final developmental component (DCs) score. See code deconvolution.R for additional details. Finally, every DC score was min-max normalized across all samples for each developmental identity and used for radar plots in Fig. 4B-E and Fig. S4–7. Additionally, these scores were used as input for the D-MLP(see below).

### Pan-cancer comparisons of tumor-normal and tumor grade and embryonic period score calculation

The embryonic period score, shown in Fig. 3, was calculated as follows. For every sample the embryonic period score was calculated by multiplying the number of the most strongly correlated cells, as described above, by their embryonic period of origin (E9.5-E13.5), as in the previously identified signature. See code Fig.3.R. For tumor-normal analysis, the TCGA tumor cohort was subset to those tumor types that only had both tumor and normal samples analyzed. For low-high grade analysis, the TCGA tumor cohort was subset to those tumor type that only had grade clinical information. A list of samples used is given in Supplementary Table 3. For categoric enrichment testing, the embryonic period of the most highly correlated cells to each sample was summed within each category to create a 2×5 grid (Tumor-Normal vs embryonic time or Grade2-Grade3 vs embryonic time) and tested using chi-squared (χ2) as (observed - expected) / sqrt(expected). Enrichment is calculated as the relative residuals from chi-squared testing and displayed in Fig. 2C and Fig. 2E.

### Identification of positive, negative, unrelated and variable associations for trajectories

Normalized correlation coefficients (NCCS) between developmental sub-trajectories and tumor samples (Fig. 2D-E) were categorized into 4 classes based on their mean and standard deviation as follows: mean _nccs < -0.5 were classified as negative, mean _nccs > 0.5 were classified as positive, sd_nccs > 0.4 were classified as variable, and the remaining relationships were considered uninformative. See also Fig. S2.

### Differential developmental component (DC) score analysis

For each tumor type the samples were categorized as concordantly or discordantly predicted based on the results of the classifier as compared to the annotated diagnosis in the TCGA, Non-TCGA, or MGH cohorts (as given by histopathological workup and clinical annotation). Concordantly and discordantly classified samples were tested for differentially represented DCs: normalized DCs (as described above) were tested for differential “expression” using the linear model coupled with the empirical Bayes statistics implemented in limma (v3.40.6) (6), Bioconductor (https://CRAN.R-project.org/package=BiocManager, v1.30.10) R (https://www.R-project.org) packages. Only DCs with a Benjamini-Hochberg adjusted p-value < 0.05 were considered to be differential between concordantly and discordantly predicted samples. See code Differential.R for further details.

### Developmental Multilayer Perceptron (D-MLP)

The Developmental Multilayer Perceptron (D-MLP) classifier has been trained to take as input the min-max normalized scores of the 214 developmental components, as is, and provide as output the likelihood score for every 27 possible tumor type labels. The final model has the following architecture: 1 input layer with 214 nodes, 2 hidden layers of 800 and 200 nodes, respectively, 1 output layer with 27 nodes. The model was compiled using stochastic gradient descent as optimizer, mean squared error as loss indicator and accuracy as metric. The model was trained on test set for 300 epochs with an early stop function monitoring accuracy score with a patience of 3. The hyper-parameters of the models for optimization include: 1) number of hidden layers, 2) the number of nodes per layer, 3) the type of optimizer function, 4) the type of loss indicator, and 5) the number of epochs. These were found via a set of grid searches; see Table S8 for additional details. The D-MLP was written in python (v3.6.4) using sklearn (v0.19.1) and keras (v2.2.0, with TensorFlow backend). See also code mlp_classifier.py.

### Benchmark classifiers and performance

The performance of the D-MLP classifier trained on the developmental components was tested against models (sharing the same architecture as described above) trained on pure gene expression profiles directly without developmental deconvolution. We opted for two sets of genes: I) Clinical oncopanel genes and II) highly variable genes. I) Clinical oncopanel genes represent a list of 251 genes tested in routine clinical cancer care at the Massachusetts General Hospital (assays: SNaPshot, Solid Fusion Assay, Heme SNaPshot); the full list is given in Table S8. Since this list contains more features (genes) than developmental components used in the D-MLP classifier, we generated 10 random subsets of 214 genes out of 251. Each of these 214 gene subsets was used to train the D-MLP classifier, and the highest performing classifier out of these 10, assessed by top1 and top3 accuracy, was directly compared against the developmental based classifier (D-MLP) as shown (Fig. 5C, Fig. S10). For the highly variable gene classifier, the expression profile of the full pan-cancer cohort (TCGA, NON-TCGA and MGH) was considered. The highly variable gene set represents the 214 most variably expressed genes assessed by pure variance (calculated by *var* function in R across the expression dataframe).

### UMAP

The Uniform Manifold Approximation and Projection for Dimension Reduction (UMAP) analysis was performed using R implementation of UMAP algorithm (48). For both UMAPs (All TCGA [Ut] tumor types and the one on lung and breast samples Ulb only, Fig. 4E-F and Fig. S10 respectively), standard parameters were used with the following exceptions: Ut: a=.5, b=.75, Ulb: a=.1, b=.75.

### Statistical analysis

Statistical analysis reported in this work were performed using R (v3.6.3). Enrichment (Fig. 3) was calculated using χ2 test and represented as (observed - expected) / sqrt(expected). Statistical differences between cumulative distributions were evaluated by means of Kolmogorov-Smirnov (K-S) tests in Figs. 3D and 3F. Pairwise differences between means of continuous variables from different samples were evaluated using Mann-Whitney tests in Figs. 3B. Receiver-operator characteristics were calculated using roc_curve and roc_auc functions from Python3 sklearn (v0.22.1) using 1,000 bootstraps to calculate empirical 95% confidence intervals. See code test_classifier.py for further details.

### Figures

Boxplots, ECDF plots, violin plots and bar plots were generated using the ggplot2 v 3.3.2 package. Detailed commands can be found in scripts Fig3.R and Fig5.R. Heatmaps were generated with pheatmap (v1.0.12) (49) package using functions described in Fig2.R and Fig6.R. UMAP plots were generated with *plot()* native R as given in the scripts for individual figures. Radar plots were generated in R using a custom function given in script Fig4.R. Graphical representations were generated using Biorender (https://biorender.com/).

### Data and code availability

Code is available at https://github.com/emoiso/DevTum. Original data is available on TCGA and MOCA websites; file intermediates generated in this study are available at (https://www.dropbox.com/sh/35dg8c5lp32kuwy/AAAir8uo4Xmz_X4lysQDWZyGa?dl=0). Code both to generate the D-MLP and completed model code is available in the above folders. It is noted here this AI algorithm cannot be used in routine practice as it is not FDA approved and is included for research and evaluation purposes. Code and intermediate data files are in the process of further deposition in public repositories.

### Code description

Correlation.R: this file contains the main commands to generate the correlation coefficients between tumor samples (TCGA, NON TCGA and MGH) and the high quality MOCA cells.

Data.recap.R: this file contains the main command to summarize and postprocess the results of the correlation analysis and organize them for further analysis.

Overall Z scores.R: this file contains the main command to normalize and scale the similarity scores Deconvolution.R: this file contains the main command for performing developmental deconvolution and associated normalization/scaling.

Fig2.R: this file contains the main command to generate the data and the plots in figures 2A-C, 3A and S1

Fig3.R this file contains the main command to generate the data behind figures 3C-F, relative bar plots, and the empirical cumulative distribution function (CDF) plots

Fig4.R this file contains the main command to generate radar plot as shown in figure 4A-E and S4-7

Fig5.R this file contains the main command for the analysis and graphical representation of the classifier results. In particular it contains the code for Sankey plots, bar plots, and ROC curve plotting in Fig. 5

Fig6.R this file contains the main commands for generating the two heatmaps and the bar plots in Fig. 6. Differential.R this file contains the main command to perform differential component analysis.

Mgh_sequencing.sh this file contains the main commands to perform alignment and gene quantification steps starting from fastq files.

Test_classifier.py this file contains the commands used to generate receiver operator curve characteristics and confidence intervals from MLP output

### Data description

Supplementary Data 1: this file contains summary statistics for the MGH samples alignment; in particular, the number of input reads, the percentage of uniquely, multi- and unmapped reads at the sample level.

Supplementary Data 2: this file contains the gene expression profile of MGH samples, measured in transcripts per million (TPM), as calculated by RSEM using the above protocols

## Supplementary Tables

Table S1: TCGA cohort samples used in this study given by TCGA sample code

Table S2: Summary statistics for correlation coefficients shown in Figure 2 by tumor type

Table S3: Pan-cancer cohorts used for analyses in Figure 3

Table S6: Gene name conversions between mouse and human samples along with TCGA gene IDs

Table S7: Conversion of non-TCGA study tumor types to codes used in D-MLP classification

Table S8: Hyperparameter optimization for D-MLP and genes used for benchmark classifiers

